# An exercise-modifiable mitochondrial basis for skeletal muscle insulin resistance in humans

**DOI:** 10.64898/2025.12.17.25342274

**Authors:** Tue Leth Nielsen, Michelle Damgaard, Nicoline Resen Andersen, Morten Dunø, Anja Lisbeth Frederiksen, Gerrit van Hall, Lykke Sylow, Marta Murgia, Steen Larsen, Signe S. Torekov, John Vissing, Matteo Fiorenza

## Abstract

Mitochondrial dysfunction has long been associated with insulin resistance (IR), yet the causal relationship in humans remains unresolved. Here, to probe causality and delineate the underlying molecular mechanisms, we leveraged carriers of the m.3243A>G mtDNA variant as a human genetic model of primary mitochondrial dysfunction linked to diabetes. Deep metabolic phenotyping revealed selective skeletal muscle IR featuring preserved insulin-stimulated Akt-TBC1D4 signaling but blunted mTORC1 activation. Integrated muscle proteomic and bioenergetic profiling demonstrated broad mitochondrial protein depletion, OXPHOS- and complex I-specific deficits, and concomitant AMPKγ2 downregulation. An exercise-based intervention improved muscle insulin sensitivity, partially rescued mitochondrial defects, and increased AMPKγ2 abundance, without altering Akt-TBC1D4 or mTORC1 signaling. An integrative framework linking phenotypic features and exercise-induced remodeling signatures confirmed rescue of m.3243A>G-related molecular alterations and their association with muscle insulin sensitivity. These findings define an exercise-modifiable mitochondrial basis for skeletal muscle IR and identify specific mitochondrial defects as candidate targets for improving glucose homeostasis in common insulin-resistant states.

## INTRODUCTION

Peripheral insulin resistance (IR) is a hallmark of type 2 diabetes mellitus (T2DM)^1^. Skeletal muscle accounts for the majority of insulin-stimulated glucose disposal^2^, hence preserving or restoring muscle insulin sensitivity is crucial for the prevention and treatment of T2DM. While current pharmacotherapies, including incretin-based agents and thiazolidinediones, can improve systemic insulin sensitivity, no existing drug directly enhances muscle insulin action^3^. This underscores the need to identify novel molecular targets for improving skeletal muscle insulin sensitivity.

Mitochondrial dysfunction, used here as a shorthand term for quantitative and qualitative impairments across multiple domains of mitochondrial biology, has long been implicated in the pathophysiology of skeletal muscle IR^4–7^. However, whether mitochondrial dysfunction represents a cause or consequence of IR has not been conclusively demonstrated^8^. Clarifying this causal relationship is critical for advancing our understanding of diabetes pathophysiology as well as the cellular consequences of mitochondrial abnormalities, thereby informing mitochondrial biology and the potential of mitochondria-targeted therapeutic strategies.

Rare genetic variants that selectively perturb a defined biological process offer a powerful approach to infer causal links between cellular mechanisms and physiological phenotypes in humans^9,10^, as their downstream consequences can reveal mechanistic relationships otherwise obscured in complex multifactorial diseases. Primary mitochondrial diseases, caused by pathogenic variants in nuclear or mitochondrial DNA genes encoding proteins governing mitochondrial function, exemplify this principle, as they are often accompanied by metabolic abnormalities^11–13^, and thereby offer a unique opportunity to dissect the causal role of mitochondrial dysfunction in common metabolic disorders.

The m.3243A>G variant in the *MT-TL1* gene, encoding mitochondrial tRNA^Leu(UUR)^, is the most common pathogenic mitochondrial DNA (mtDNA) variant^14^ and is associated with a markedly higher prevalence and penetrance of diabetes than other mtDNA variants^15,16^. These features make individuals carrying m.3243A>G (hereinafter “carriers”) a particularly informative model for investigating how mitochondrial dysfunction contributes to impaired glucose homeostasis in humans. Importantly, although the diabetic phenotype associated with m.3243A>G has been attributed to both IR and pancreatic β-cell dysfunction^17,18^, the relative contribution of each defect and the molecular mechanisms involved remain incompletely defined^19–22^. In particular, whether m.3243A>G-associated IR is tissue-specific and which mitochondrial alterations underlie it, especially in skeletal muscle given its high mitochondrial density, have not been established.

Taken together, despite extensive research linking mitochondrial dysfunction to IR, whether mitochondrial derangements directly impair insulin action in humans remains unresolved. Prior investigations in insulin-resistant individuals^6,7,23–26^ and in human models of acute mitochondrial stress^27–29^ have provided valuable insight but have been unable to establish causality or capture the consequences of chronic mitochondrial dysfunction on insulin action. Moreover, the specific mitochondrial defects and the molecular pathways through which they compromise insulin sensitivity remain poorly defined. In this context, m.3243A>G carriers provide a unique human genetic model in which chronic mitochondrial dysfunction is the primary defect, enabling direct characterization of the mitochondrial abnormalities underlying impaired insulin action; an insight unattainable in the more heterogeneous forms of IR that characterize T2DM.

To directly interrogate the mechanistic link between mitochondrial dysfunction and IR in humans, we conducted two complementary physiological studies in m.3243A>G carriers. First, in a cross-sectional study integrating comprehensive *in vivo* metabolic phenotyping with skeletal muscle molecular and bioenergetic profiling, we defined the mitochondrial defects associated with the insulin-resistant phenotype of m.3243A>G carriers and interrogated candidate molecular mechanisms linking these abnormalities to impaired muscle insulin action. Second, in an exercise-based intervention study, we tested whether improvements in muscle insulin sensitivity coincide with rescue of m.3243A>G-associated mitochondrial defects, thereby providing convergent evidence supporting a causal contribution of mitochondrial dysfunction to muscle IR. Together, this integrative defect-rescue framework identified distinct, exercise-modifiable mitochondrial signatures of muscle IR and delineated molecular features linking mitochondrial dysfunction to impaired insulin sensitivity in humans.

## RESULTS

### Participant characteristics

In a cross-sectional phenotyping study, we compared metabolic, molecular, and bioenergetic profiles between fifteen m.3243A>G carriers and fifteen healthy controls individually matched for age, sex, body mass index (BMI), and objectively measured habitual physical activity (Fig. 1A and Table S1).

**Figure 1.**
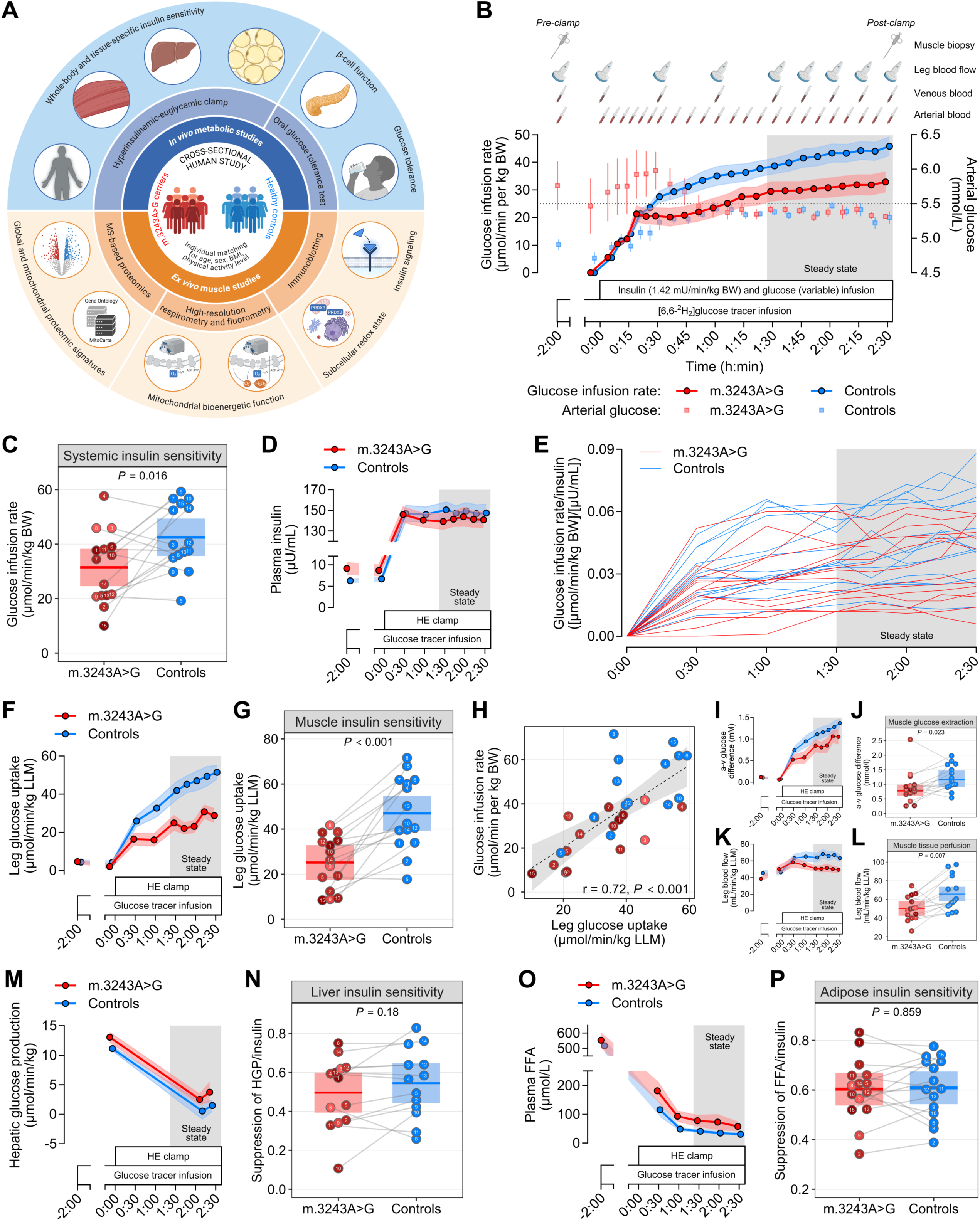
Skeletal muscle is the principal site of insulin resistance in m.3243A>G carriers. (A) Schematic overview of the cross-sectional study design. (B) Workflow of the hyperinsulinemic-euglycemic (HE) clamp visit, including the mean glucose infusion rate corrected per body weight (BW) and the arterial glucose concentration. The dotted line indicates arterial glucose concentration corresponding to euglycemia. Data presented as means ± SEM. (C) Whole-body insulin sensitivity expressed as mean glucose infusion rate during the steady-state period of the HE clamp. (D) Time course of plasma insulin levels (data are means ± SEM). (E) Interindividual and time-dependent variability of the glucose infusion rate corrected per plasma insulin during the HE clamp. (F and G) Skeletal muscle insulin sensitivity expressed as the leg glucose uptake, as calculated from the arteriovenous (a-v) difference in plasma glucose concentration and the femoral arterial blood flow corrected per leg lean mass (LLM). [(F)] time course of leg glucose uptake (data are means ± SEM). [(G)] leg glucose uptake during the steady-state period of the HE clamp. (H) Pearson’s correlation between muscle (leg glucose uptake) and whole-body (glucose infusion rate) insulin sensitivity. (I to L) Arteriovenous (a-v) difference in plasma glucose concentration and femoral arterial blood flow corrected per leg lean mass (LLM). [(I) and (K)] time course of a-v glucose difference and leg blood flow (data are means ± SEM). [(J) and (L)] a-v glucose difference and leg blood flow during the steady-state period of the HE clamp. (M and N) Liver insulin sensitivity expressed as the insulin-dependent suppression of hepatic glucose production, as calculated from the difference in hepatic glucose production between the basal state and the steady-state period of the HE clamp divided by plasma insulin levels during the steady-state period. [(M)] time course of hepatic glucose production (data are means ± SEM). [(N)] Suppression of hepatic glucose production (HGP) corrected for plasma insulin levels. (O and P) Adipose tissue insulin sensitivity expressed as the insulin-dependent suppression of lipolysis, as calculated from the difference in plasma free fatty acids (FFA) between the basal state and the steady-state period of the HE clamp divided by plasma insulin levels during the steady-state period. [(O)] time course of plasma FFA (data are means ± SEM). [(P)] Suppression of plasma FFA corrected for plasma insulin levels. Linear mixed models were used to estimate between-group differences [(C), (G), (J), (L), (N) and (P)]. Data are presented as observed individual values (with lines connecting individually matched participants) and estimated means ± 95% confidence limits, unless otherwise stated. For m.3243A>G carriers, individual datapoints are color-shaded to indicate muscle mtDNA heteroplasmy (light red = low, dark red = high). n = 30. Illustrations in (A) were created with BioRender.com.

Fasting plasma glucose, insulin, C-peptide, HbA1c, and HOMA2-IR were elevated in carriers, consistent with the established association between the m.3243A>G variant and impaired glucose homeostasis^17,19,21^. Cardiorespiratory fitness (V̇O_2_peak) was reduced in carriers compared to controls, despite similar physical activity levels, pointing to intrinsic limitations in oxygen transport and utilization^30^. Among the carriers, skeletal muscle mtDNA heteroplasmy (ratio of mutated-to-wild type mtDNA) spanned 1-98% (Fig. S1A and Table S2). Five carriers were clinically asymptomatic, while the remaining carriers presented with at least one clinical feature of mitochondrial disease (Table S2). Two carriers had established diabetes diagnosis and were receiving glucose-lowering medications upon enrolment in the study. An additional four carriers without known diabetes met diagnostic criteria for diabetes^31^ during the study.

### Skeletal muscle is the principal site of insulin resistance in m.3243A>G carriers

To assess systemic and tissue-specific insulin sensitivity, we combined hyperinsulinemic-euglycemic (HE) clamps with the femoral arteriovenous balance technique and glucose tracer infusion (Fig. 1B). Whole-body insulin sensitivity, quantified by the glucose infusion rate required to maintain euglycemia, was 26% lower in m.3243A>G carriers compared to controls (Fig. 1C). Plasma insulin remained comparably elevated throughout the HE clamp (Fig. 1D), excluding differences in circulating insulin as a cause of the observed heterogeneity in systemic insulin sensitivity (Fig. 1E).

Skeletal muscle insulin sensitivity, determined from the net plasma glucose exchange across the leg, was reduced by 46% in carriers (Fig. 1F and G) and showed a strong positive correlation with whole-body insulin sensitivity (Fig. 1H). The impairment in muscle insulin sensitivity resulted from both diminished arteriovenous glucose difference, reflecting reduced glucose extraction, and decreased leg blood flow, indicating reduced muscle tissue perfusion (Fig. 1I to L).

To determine whether other insulin-sensitive tissues contributed to systemic IR, we evaluated liver- and adipose-specific insulin sensitivity. Insulin-mediated suppression of endogenous glucose production (Fig. 1M and N) and plasma free fatty acids (Fig. 1O and P) were comparable between carriers and controls, demonstrating preserved liver and adipose tissue insulin action.

Given the marked reduction in insulin-stimulated muscle glucose uptake, we next examined whether muscle IR resulted in differences in muscle substrate oxidation using leg-specific indirect calorimetry (Fig. S2A). Under hyperinsulinemia, the leg respiratory quotient was comparable between carriers and controls (Fig. S2B), indicating no shift in oxidative substrate preference. Consistent with the lower insulin-stimulated muscle glucose uptake, carriers exhibited 41% lower carbohydrate oxidation with unaltered fatty acid oxidation (Fig. S2C) and 43% lower net lactate release (Fig. S2D-E), reflecting reduced glucose flux through both oxidative and non-oxidative pathways.

Collectively, these findings demonstrate a muscle insulin-resistant phenotype in m.3243A>G carriers, while liver and adipose tissue insulin sensitivity remain preserved, suggesting skeletal muscle as the principal determinant of systemic IR.

### Impaired glucose tolerance due to concomitant defects in β-cell function and systemic insulin sensitivity in m.3243A>G carriers

To further characterize factors contributing to glucose homeostasis, we assessed glucose tolerance and pancreatic β-cell function during an oral glucose tolerance test (OGTT) (Fig. S3A). Carriers displayed impaired glucose tolerance, as measured by a 2.6-fold higher incremental area under the curve (iAUC) of glucose (Fig. S3B-C), in line with previous observations^18,19,21,32^. Both early-phase insulin secretion capacity, assessed by the insulinogenic index, and the OGTT-based insulin sensitivity index, estimated by the Matsuda index, were reduced in carriers (Fig. S3D-F). This combined defect resulted in a 65% lower pancreatic β-cell function, quantified by the disposition index, compared with controls (Fig. S3G). This finding aligns with one prior study^17^ but contrasts with reports of preserved β-cell function in other m.3243A>G cohorts^19–21^, likely reflecting the heterogeneous manifestations of the variant due to variable mtDNA heteroplasmy across individuals. To address this, we examined associations between mtDNA heteroplasmy and parameters of glucose homeostasis, which revealed that only the insulinogenic index and the disposition index were inversely associated with mtDNA heteroplasmy (Fig. S4A). OGTT-based indexes of liver and adipose tissue insulin sensitivity were comparable in carriers and controls (Fig. S3H-J), thus supporting the preserved liver and adipose insulin action resulting from the HE clamp.

Taken together, m.3243A>G carriers exhibit impaired glucose tolerance driven by concomitant defects in pancreatic β-cell function and systemic insulin sensitivity, which together contribute to their elevated risk of developing diabetes.

### Impaired insulin-stimulated mTORC1 activation accompanies muscle insulin resistance in m.3243A>G carriers

To explore molecular mechanisms underlying muscle IR in m.3243A>G carriers, we examined phosphorylation of key proteins modulating the insulin signaling pathway in skeletal muscle (Fig. 2A). Insulin-stimulated phosphorylation of both proximal (Akt2) and distal (GSK3β and TBC1D4) nodes did not differ between carriers and controls (Fig. 2A-D), partly consistent with the preserved proximal insulin signaling reported in common IR^33,34^. By contrast, basal (non-insulin-stimulated) Akt^Ser473^ phosphorylation was reduced in carriers, aligning with recent phosphoproteomics data from insulin-resistant individuals^35^ and likely reflecting chronic hyperinsulinemia-driven Akt desensitization^36^. Given emerging evidence that human muscle IR is primarily characterized by signaling defects beyond the canonical Akt-TBC1D4 axis^35,37^, we next examined mTORC1 and AMPK signaling. Insulin-stimulated phosphorylation of the mTORC1 effector p70S6K was reduced in carriers (Fig. 2E). In contrast, insulin-stimulated AMPKα2^Thr172^ phosphorylation was comparable in carriers and controls (Fig. 2F).

**Figure 2.**
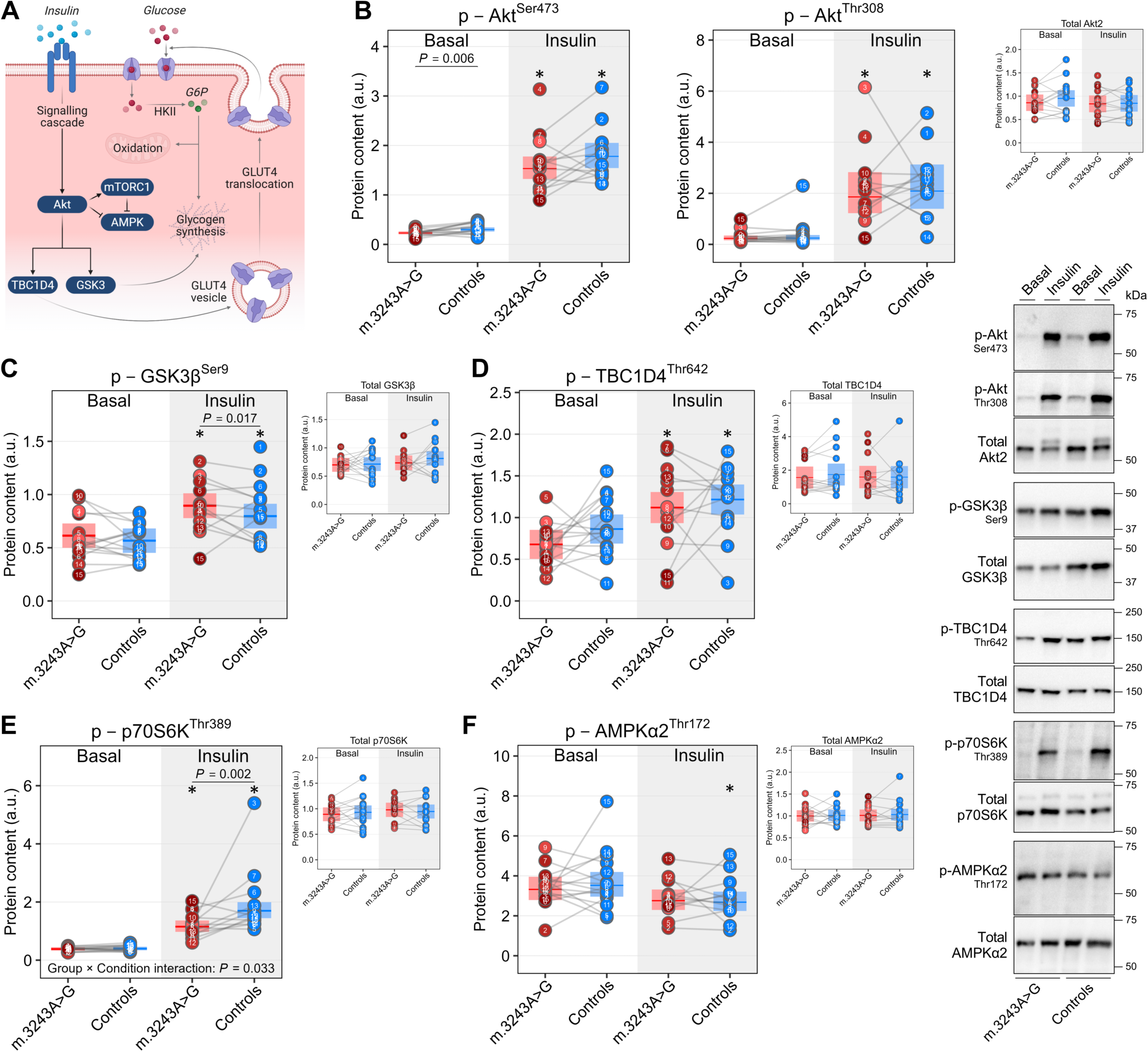
Impaired insulin-stimulated mTORC1 activation accompanies muscle insulin resistance in m.3243A>G carriers. (A) Schematic overview of key signaling nodes modulating muscle insulin action. (B to D) Canonical insulin signaling as determined by phosphorylation of Akt2 on Ser473 and Thr308, and phosphorylation of the Akt substrates GSK3β on Ser9 and TBC1D4 on Thr 642 in whole-muscle lysates from skeletal muscle biopsy samples obtained before (Basal) and immediately after (Insulin) the hyperinsulinemic-euglycemic (HE) clamp. Levels of total Akt2, GSK3β, and TBC1D4 were comparable across groups and unaffected by insulin. (E and F) mTORC1 and AMPK signaling as determined by phosphorylation of p70 S6 kinase (p70S6K) on Thr389 and AMPKα2 on Thr172 in whole-muscle lysates from skeletal muscle biopsy samples obtained before (Basal) and immediately after (Insulin) the HE clamp. Levels of total p70S6K and AMPKα2 were comparable across groups and unaffected by insulin. Linear mixed models were used to estimate within- and between-group differences. Data are presented as observed individual values (with lines connecting individually matched participants) and estimated means ± 95% confidence limits, unless otherwise stated. For m.3243A>G carriers, individual datapoints are color-shaded to indicate muscle mtDNA heteroplasmy (light red = low, dark red = high). *Different from Basal (*P* < 0.05). Basal, n = 30; Insulin, n = 27 (13 in m.3243A>G, 14 in Controls). Illustrations in (A) were created with BioRender.com.

Collectively, these results indicate that skeletal muscle IR in m.3243A>G carriers manifests despite preserved insulin signaling through the canonical Akt-TBC1D4 axis and is accompanied by reduced basal Akt activity and diminished insulin-stimulated mTORC1 activation.

### Global muscle proteomic signatures of m.3243A>G carriers

To uncover molecular signatures of muscle IR in m.3243A>G carriers, we employed a high-throughput mass spectrometry-based proteomics workflow on skeletal muscle biopsies (Fig. 3A; one carrier excluded for insufficient tissue), which quantified over 4,000 proteins across all samples (range: 4,231-4,803; Fig. 3B). Ranking plots showed that proteins with the highest and lowest abundances were similarly ranked between carriers and controls (Fig. 3C), and principal component analysis revealed substantial heterogeneity on PC1 that was not primarily driven by the m.3243A>G genotype (Fig. 3D), together indicating preservation of the core skeletal muscle proteome and absence of a broad global proteomic shift associated with m.3243A>G . Comparative analyses identified 507 differentially abundant proteins between carriers and controls (288 downregulated and 219 upregulated; unadjusted *P* < 0.05), of which 9 were significantly downregulated and 12 significantly upregulated in carriers after multiple testing correction (false discovery rate [FDR] < 0.05) (Fig. 3E). The most significantly downregulated proteins included regulators of cellular energy metabolism such as AMP-activated protein kinase subunit gamma-2 (PRKAG2) and mitochondrial coenzyme A diphosphatase (NUDT8) (Fig. 3F). Additional downregulated proteins (CKMT1B, PPP1R3B, RMDN2) converged on phosphocreatine turnover, glycogen synthesis, and possibly on microtubule-mediated GLUT4 trafficking^38^, while upregulated RXRB implicates PPAR/RAR signaling and myogenic and metabolic transcriptional programming in muscle^39^. Core insulin action proteins (INSR, GLUT4, hexokinases) were unaltered in carriers (Fig. 3G).

**Figure 3.**
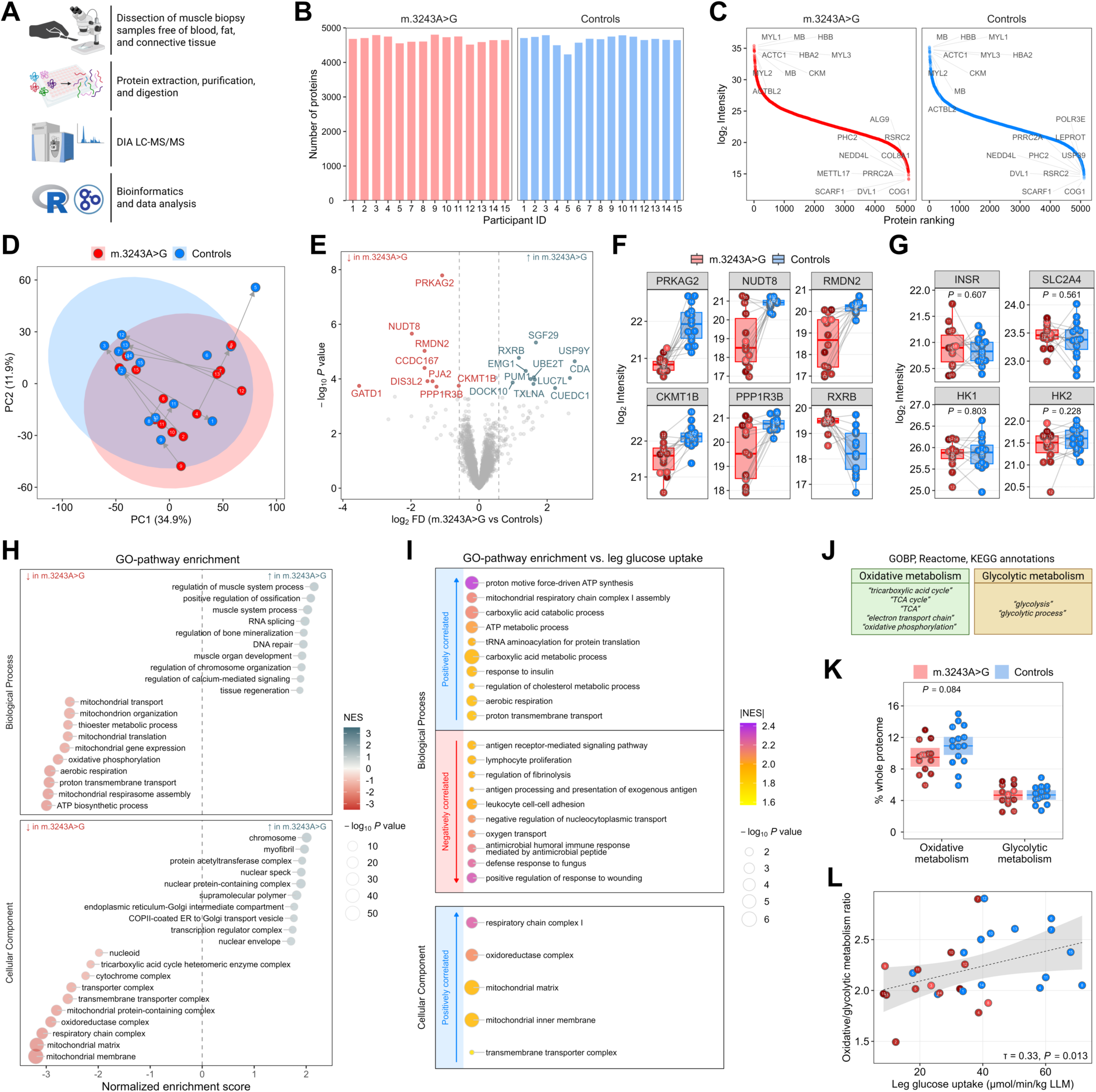
Global muscle proteomics signatures of m.3243A>G carriers. (A) Schematic overview of the mass spectrometry-based muscle proteomics workflow. (B) Number of muscle proteins quantified per participant in each group. (C) Dynamic range plot of ranked protein abundance (log_2_ intensity) showing the top and bottom ten ranked proteins. (D) Principal component (PC) analysis of the muscle proteome. (E) Volcano plot of differentially abundant muscle proteins in m.3243A>G carriers vs. controls. Downregulated and upregulated proteins in m.3243A>G carriers are marked in red and blue, respectively (FDR < 5%). (F) Boxplots of selected muscle proteins related to metabolism and differentially abundant in m.3243A>G carriers compared to controls. (G) Abundance of insulin receptor (INSR), GLUT4 (SLC2A4), hexokinase I (HK1), and hexokinase II (HK2) in m.3243A>G carriers compared to controls (unadjusted *P* values are reported). (H) Bubble plots of most significantly depleted and enriched Gene Ontology (GO) Biological Process and Cellular Component terms in m.3243A>G carriers. Bubble plots are colored by normalized enrichment score, and size is based on *P_adj_*. (I) Within-pair GO-pathway enrichment based on Kendall’s rank correlation with insulin-stimulated leg glucose uptake in positive and negative directions. Bubble plots are colored by absolute normalized enrichment score, and size is based on *P_adj_*. (J) GOBP, Reactome, and KEGG annotation terms defining oxidative and glycolytic metabolism proteins. (K) Proportion of oxidative and glycolytic metabolism proteins relative to the whole proteome. (L) Kendall’s rank correlation of muscle insulin sensitivity (leg glucose uptake during the hyperinsulinemic-euglycemic clamp) with the ratio of oxidative/glycolytic metabolism protein abundance. Linear mixed models were used to estimate between-group differences (K). Data are presented as observed individual values (with lines connecting individually matched participants) and estimated means ± 95% confidence limits, unless otherwise stated. For m.3243A>G carriers, individual datapoints are color-shaded to indicate muscle mtDNA heteroplasmy (light red = low, dark red = high). n = 29 (14 in m.3243A>G, 15 in Controls). Illustrations in (A) and (I) were created with BioRender.com.

To gain insight into m.3243A>G-dependent alterations in biological processes and cellular components, we performed Gene Ontology (GO)-based pathway enrichment analysis, which revealed marked depletion of mitochondrial oxidative and metabolic processes and corresponding cellular components, in contrast to enrichment of myogenic and contractile pathways in carriers (Fig. 3H). To interrogate proteomic determinants of muscle insulin sensitivity, we conducted within-pair rank correlation with insulin-stimulated leg glucose uptake, revealing that enrichment of mitochondrial oxidative processes and compartments, most notably mitochondrial complex I, was positively associated with greater muscle insulin sensitivity (Fig. 3I).

We next compared the relative abundance of oxidative versus glycolytic metabolism proteins (Fig. 3J), which revealed a tendency towards selective reduction of oxidative, but not glycolytic, proteins in carriers (Fig. 3K). The resulting oxidative-to-glycolytic proteome ratio was positively associated with muscle insulin sensitivity (Fig. 3L), consistent with prior evidence linking this ratio to whole-body insulin sensitivity^35^.

Taken together, skeletal muscle proteomic profiling reveals coordinated downregulation of key metabolic proteins together with depletion of mitochondrial pathways in m.3243A>G carriers, highlighting disrupted oxidative metabolism as a key feature of their insulin-resistant phenotype.

### Selective remodeling of the muscle mitochondrial proteome in m.3243A>G carriers

Given the pronounced alterations in mitochondrial and oxidative processes, we next conducted a targeted analysis of the mitochondrial proteome using the MitoCarta3.0 database^40^ (Fig. 4A). Mitochondrial protein enrichment, measured as the summed intensity of mitochondrial proteins relative to the total proteome, was significantly lower in carriers (Fig. 4B), indicating reduced mitochondrial content in m.3243A>G skeletal muscle. Enrichment analysis based on MitoPathways from MitoCarta3.0 revealed marked depletion of pathways related to oxidative phosphorylation (OXPHOS) and substrate metabolism in carriers, with complex I subunits, mitochondrial ribosome, and TCA cycle proteins resulting among the most depleted (Fig. 4C).

**Figure 4.**
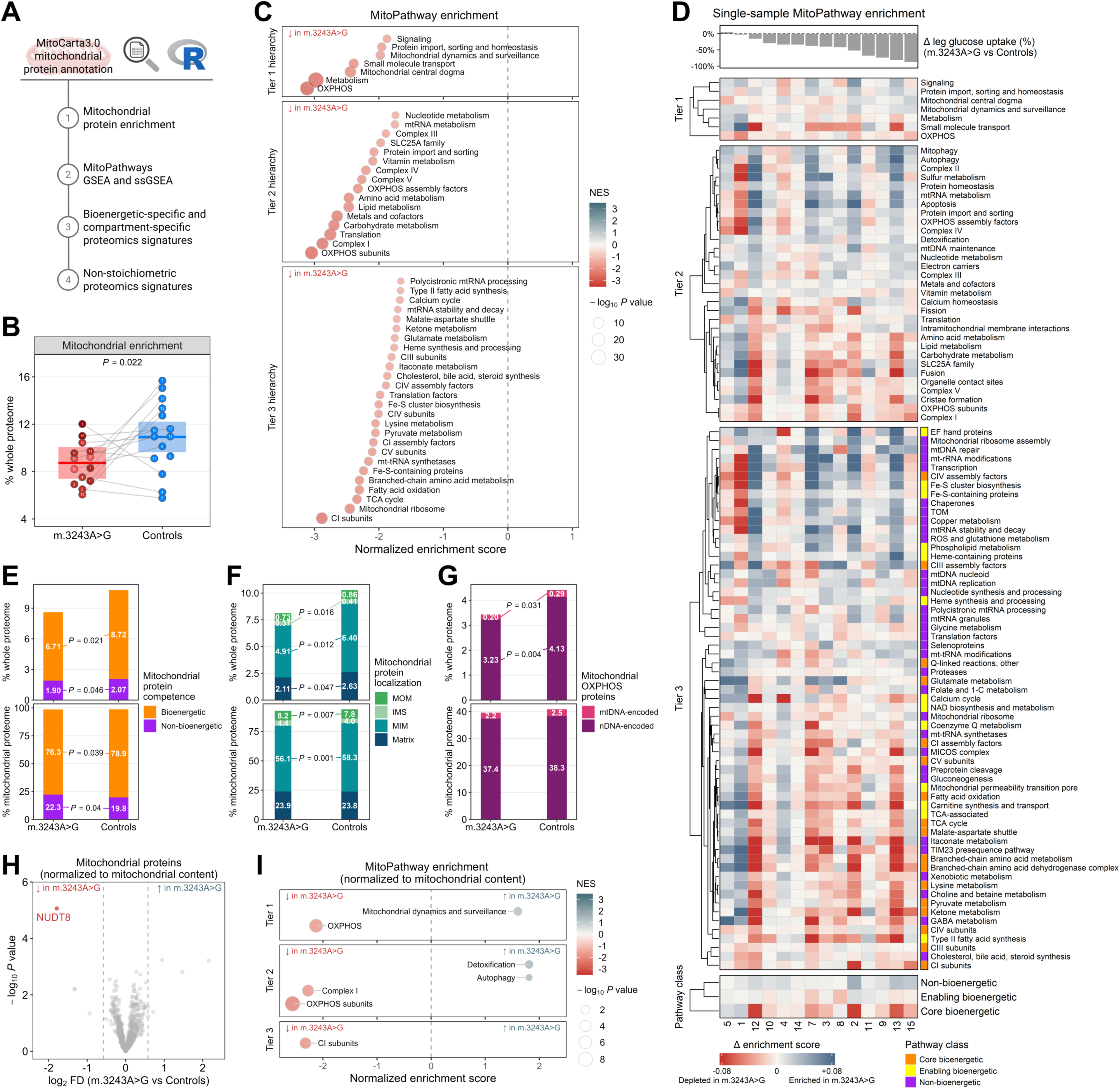
Selective remodeling of the muscle mitochondrial proteome in m.3243A>G carriers. (A) Schematic overview of the mitochondrial proteomics analytical workflow. (B) Mitochondrial protein enrichment factor as determined by the summed mitochondrial protein abundance relative to the whole proteome. (C) Bubble plot showing significantly depleted and enriched mitochondrial pathways (MitoPathways from MitoCarta3.0) in m.3243A>G carriers. Bubble plots are colored by normalized enrichment score and size is based on *P_adj_*. (D) Hierarchically clustered heatmap showing matched carrier-control differences in MitoPathways enrichment scores. Tier 3 MitoPathways are color-coded by functional class (*core bioenergetic*, *enabling bioenergetic*, *non-bioenergetic*). The top bar plot indicates the corresponding difference in muscle insulin sensitivity (leg glucose uptake during the hyperinsulinemic-euglycemic clamp), while the lower panel summarizes enrichment across pathway classes. (E) Proportion of bioenergetically (“*core bioenergetic*” and “*enabling bioenergetic*” annotated proteins) and non-bioenergetically competent mitochondrial protein abundance relative to the whole and mitochondrial proteome. (F) Proportion of compartment-specific mitochondrial protein abundance relative to the whole and mitochondrial proteome. MOM, mitochondrial outer membrane; IMS, intermembrane space; MIM, mitochondrial inner membrane. (G) Proportion of mitochondrial oxidative phosphorylation (OXPHOS) system proteins (complex I, II, III, IV, V) encoded by either mitochondrial DNA (mtDNA) or nuclear DNA (nDNA) relative to the whole and mitochondrial proteome. (H) Volcano plot of differentially abundant mitochondrial proteins in m.3243A>G carriers vs. controls after normalization to mitochondrial protein content. () Significantly enriched (blue) and depleted (red) MitoPathways in m.3243A>G carriers after normalization to mitochondrial protein content. Linear mixed models were used to estimate between-group differences [(B), (E), (F), and (G)]. Data are presented as observed individual values (with lines connecting individually matched participants) and estimated means ± 95% confidence limits. For m.3243A>G carriers, individual datapoints are color-shaded to indicate muscle mtDNA heteroplasmy (light red = low, dark red = high). n = 29 (14 in m.3243A>G, 15 in Controls). Illustrations in (A) were created with BioRender.com.

To examine how pathway-level alterations within the mitochondrial proteome relate to muscle IR, we performed single-sample enrichment analysis across MitoPathways. This approach enabled visualization of individually matched carrier-control differences in mitochondrial pathway enrichment in relation to within-pair differences in insulin-stimulated leg glucose uptake. Hierarchical clustering of single-sample enrichment profiles revealed a coordinated depletion of pathways governing mitochondrial bioenergetics in m.3243A>G carriers (Fig. 4D). When tier 3 MitoPathways were grouped into functional classes (i.e. *core bioenergetic*, *enabling bioenergetic*, and *non-bioenergetic*), the *core bioenergetic* signature showed the most pronounced reduction, whereas the *non-bioenergetic* signature was relatively enriched in m.3243A>G muscle (Fig. 4D, bottom).

To further assess whether the m.3243A>G variant differentially affects bioenergetic versus non-bioenergetic pathways, we quantified the summed abundance of proteins within each functional class. When expressed relative to the total muscle proteome, both bioenergetic and non-bioenergetic mitochondrial protein pools were reduced in carriers. In contrast, when expressed relative to the mitochondrial proteome, bioenergetically competent proteins were selectively downregulated, whereas non-bioenergetic proteins were upregulated in m.3243A>G muscle (Fig. 4E). A similar pattern was apparent across mitochondrial compartments, with proteins localized to the outer mitochondrial membrane being decreased relative to the total proteome but increased when expressed relative to the mitochondrial proteome (Fig. 4F). As m.3243A>G resides in mtDNA, we next examined whether OXPHOS proteins encoded by mtDNA were preferentially affected. mtDNA- and nuclear DNA (nDNA)-encoded OXPHOS proteins were reduced to similar extents (Fig. 4G), indicating global rather than mtDNA-selective OXPHOS protein depletion.

To determine whether the mitochondrial proteome depletion observed in carriers reflected a mere reduction in mitochondrial protein content or true remodeling of the mitochondrial proteome, we employed a normalization strategy that removes the bias introduced by differences in mitochondrial content^41,42^. Comparative analysis of the muscle mitochondrial proteome after normalization to mitochondrial protein content identified 97 proteins with differential abundance in carriers (41 downregulated and 56 upregulated; unadjusted *P* < 0.05), with only NUDT8 remaining significantly downregulated after multiple testing correction (FDR < 0.05) (Fig. 4H). MitoPathways-based enrichment analysis after normalization to mitochondrial content further revealed that OXPHOS pathways, particularly complex I, were selectively depleted, while detoxification and autophagy pathways were enriched in carriers (Fig. 4I).

Collectively, these findings indicate that mitochondrial proteome alterations in m.3243A>G carriers not only reflect a reduction in mitochondrial protein abundance but also selective remodeling of mitochondrial protein composition. Bioenergetically competent proteins were disproportionately downregulated, whereas non-bioenergetic proteins were relatively upregulated, suggesting qualitative remodeling beyond a simple loss of mitochondrial content. The persistent depletion of OXPHOS pathways, especially complex I subunits, even after normalization to mitochondrial content highlight non-stoichiometric alterations in the muscle mitochondrial proteome characterizing the insulin-resistant phenotype of m.3243A>G carriers.

### Preserved intrinsic OXPHOS capacity but selective complex I functional impairments in m.3243A>G skeletal muscle

To gain further mechanistic insight into the mitochondrial functional consequences of the m.3243A>G variant, we performed an in-depth characterization of mitochondrial bioenergetics using high-resolution respirometry and fluorometry in permeabilized muscle fibers and isolated mitochondria obtained from muscle biopsies (Fig. 5A).

**Figure 5.**
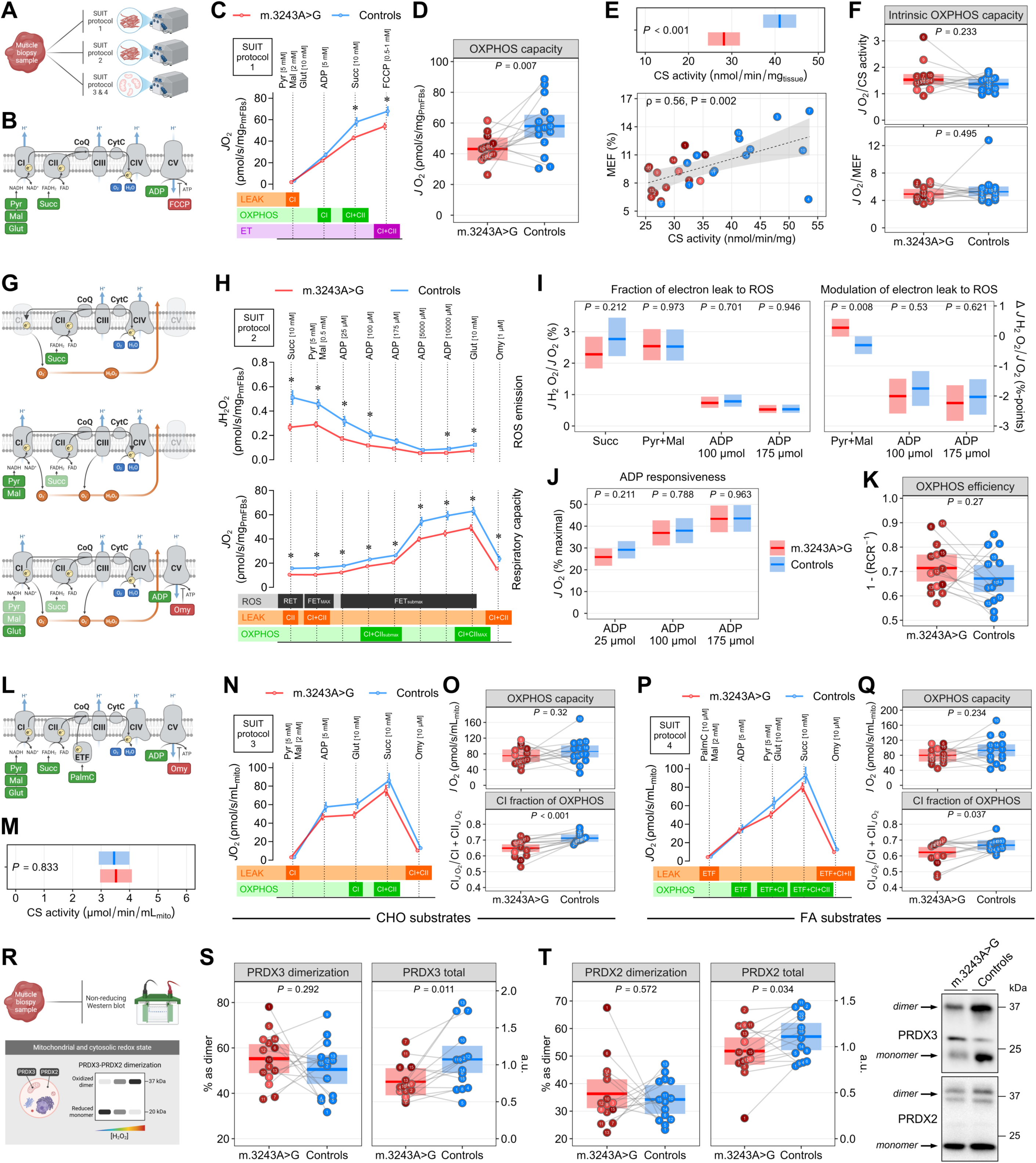
Preserved intrinsic OXPHOS capacity but selective complex I functional impairments in m.3243A>G skeletal muscle. (A) Schematic overview of the high-resolution respirometry-based muscle mitochondrial bioenergetics workflow. (B and C) Substrate-inhibitor titration (SUIT) protocol 1 used to measure *ex vivo* mitochondrial respiratory capacity (O_2_ consumption rate, *J*O_2_) in permeabilized fiber bundles (PmFBs) (data in B are means ± SEM). (D) Maximal oxidative phosphorylation (OXPHOS) capacity in PmFBs as determined by *J*O_2_ in the presence of saturating concentrations of pyruvate, malate, glutamate, ADP, and succinate. (E) Citrate synthase (CS) activity in whole muscle homogenates (reflecting mitochondrial content in PmFBs), and Spearman correlation between CS activity and the proteomics-based mitochondrial enrichment factor (MEF; related to Figure 6B). (F) Intrinsic OXPHOS capacity as determined by the maximal OXPHOS capacity normalized to readouts of mitochondrial content (CS activity and MEF). (G and H) SUIT protocol 2 used to simultaneously measure *ex vivo* mitochondrial respiratory capacity and ROS emission (H_2_O_2_ emission rate, *J*H_2_O_2_) in PmFBs (data in G are means ± SEM). (I) Fraction of electron leak to ROS as determined by the ratio between mitochondrial H_2_O_2_ emission and O_2_ consumption rates, and modulation of electron leak to ROS by pyruvate/malate and ADP. (J) Responsiveness of mitochondrial O_2_ consumption rate to submaximal ADP levels reflecting human skeletal muscle at rest. (K) Mitochondrial OXPHOS efficiency calculated as 1 − RCR = 1 − LEAK_Omy_/OXPHOS^117^. (L, N, and P) SUIT protocol 3 and 4 used to measure *in vitro* mitochondrial respiratory capacity in isolated mitochondria in the presence of either carbohydrate (CHO) or fatty acid (FA) substrate (data in L and N are means ± SEM). (M) Citrate synthase (CS) activity in isolated mitochondrial fractions. (O) Maximal OXPHOS capacity and complex I-specific contribution to OXPHOS in isolated mitochondria as determined by *J*O_2_ in the presence of CHO substrates. (Q) Maximal OXPHOS capacity and complex I-specific contribution to OXPHOS in isolated mitochondria as determined by *J*O_2_ in the presence of FA substrates. (R) Workflow to quantitatively determine the subcellular redox state in skeletal muscle biopsy samples. (S and T) Mitochondrial and cytosolic redox state as determined by protein abundance of peroxiredoxin 3 (PRDX3) and peroxiredoxin 2 (PRDX2) dimers relative to monomers. Linear mixed models were used to estimate between-group differences. Data are presented as observed individual values (with lines connecting individually matched participants) and estimated means ± 95% confidence limits, unless otherwise stated. For m.3243A>G carriers, individual datapoints are color-shaded to indicate muscle mtDNA heteroplasmy (light red = low, dark red = high). n = 30. Illustrations in (A, B, G, L, and R) were created with BioRender.com.

First, we assessed features of mitochondrial respiratory capacity in permeabilized muscle fibers (PmFBs) (Fig. 5B and C), an approach that preserves the muscle intracellular environment and more closely resembles *in vivo* mitochondrial physiology^43^. Both OXPHOS and electron transfer (ET) capacities were lower in PmFBs from m.3243A>G carriers (Fig. 5C and D). Since mitochondrial content is a key determinant of mass-specific respiratory capacity, we used citrate synthase (CS) activity as a readout of mitochondrial content^44^. CS activity was lower in carriers and correlated positively with the proteomics-derived mitochondrial enrichment factor (Fig. 5E), further supporting its validity as a marker of mitochondrial content. When OXPHOS capacity was normalized to either CS activity or proteomics-derived mitochondrial enrichment, the apparent respiratory defects were abolished (Fig 5F), indicating that reduced mitochondrial content, rather than impaired intrinsic respiratory capacity, accounts for the lower OXPHOS capacity in carriers.

Given the established link between mitochondrial oxidative stress and IR^29,45^, we next conducted simultaneous quantifications of mitochondrial ROS emission and O_2_ consumption in PmFBs (Fig. 5G and H). Absolute H_2_O_2_ emission rates were lower in carriers (Fig. 5H), but normalization to the corresponding O_2_ consumption rates (reflecting the fraction of electron leak to ROS) revealed no overall alterations in carriers (Fig. 5I), which contrasts with the elevated mitochondrial ROS production and oxidative stress reported in fibroblasts derived from m.3243A>G carriers with severe MELAS phenotypes^46–48^. However, when examining substrate-specific responses, carriers exhibited a greater increase in electron leak to ROS upon stimulation with pyruvate and malate (Fig. 5I), suggesting impaired complex I function characterized by increased electron leakage during NADH-linked substrate oxidation. Measures of ADP sensitivity and OXPHOS efficiency were unaltered in carriers (Fig. 5J and K).

To further evaluate intrinsic bioenergetic defects and uncover substrate-specific alterations, we analyzed respiratory capacity in isolated mitochondria using carbohydrate- and fatty acid-derived substrates (Fig. 5L). These experiments controlled for differences in mitochondrial content between carriers and controls, as equal mitochondrial volumes with comparable CS activity were analyzed across groups (Fig. 5M). Consistent with findings in PmFBs, isolated mitochondria from carriers and controls exhibited similar intrinsic OXPHOS capacities regardless of substrate type (Fig. 5N-Q). However, the ratio of complex I-linked to maximal (complex I+II-linked) OXPHOS capacity was lower in carriers (Fig. 5O and Q), indicating a reduced contribution of complex I to overall OXPHOS and thereby a lower reliance on NADH-linked substrate oxidation.

Because mitochondrial ROS emission rates do not necessarily reflect the intramyocellular oxidative burden that contributes to IR, we next assessed muscle mitochondrial and cytosolic redox state by measuring peroxiredoxin 3 (PRDX3) and peroxiredoxin 2 (PRDX2) dimerization (Fig. 5R)^29^. The relative abundance of PRDX3 and PRDX2 dimers was similar between carriers and controls (Fig. 5S and T), while total PRDX3 and PRDX2 protein levels (summed dimer and monomer abundance) were lower in carriers, consistent with an adaptive response to the observed reduction in mitochondrial ROS emission.

None of the measures of mitochondrial content, bioenergetics, and redox state correlated with muscle mtDNA heteroplasmy (Fig. S4B).

Together, these results demonstrate lower mass-specific OXPHOS capacity in m.3243A>G carriers, while intrinsic mitochondrial respiratory capacity and redox state remain largely preserved. However, the greater increase in electron leak to ROS during NADH-linked respiration and the diminished complex I contribution to OXPHOS reflect selective complex I functional defects that are masked under conditions of maximal respiratory flux.

### Exercise training enhances muscle insulin sensitivity in m.3243A>G carriers

To determine whether amelioration of muscle IR in m.3243A>G carriers is accompanied by partial rescue of m.3243A>G-associated mitochondrial defects, a cohort of m.3243A>G carriers (Table S1) underwent a unilateral leg cycling-based high-intensity interval training (HIIT) intervention followed by HE clamp-based assessments of muscle insulin sensitivity and muscle molecular profiling (Fig. 6A-B). Skeletal muscle mtDNA heteroplasmy was comparable across the non-trained and trained legs (Fig. S1B).

**Figure 6.**
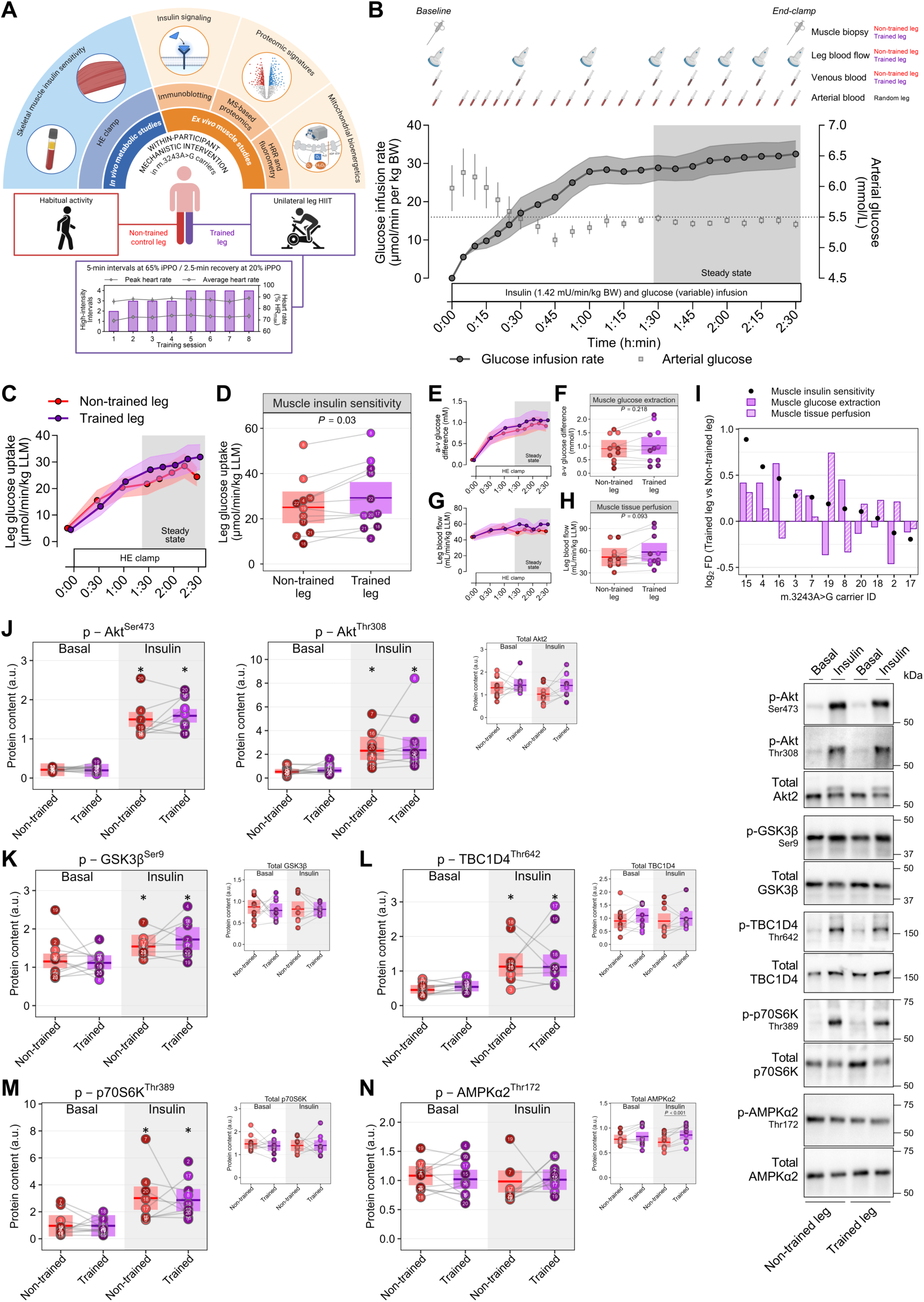
Exercise training enhances muscle insulin sensitivity in m.3243A>G carriers. (A) Schematic overview of the exercise intervention study design. (B) Workflow of the hyperinsulinemic-euglycemic (HE) clamp trial, including the mean glucose infusion rate corrected per body weight (BW) and the arterial glucose concentration. The dotted line indicates arterial glucose concentration corresponding to euglycemia. Data presented as means ± SEM. (C and D) Skeletal muscle insulin sensitivity expressed as the leg glucose uptake, as calculated from the arteriovenous (a-v) difference in plasma glucose concentration and the femoral arterial blood flow corrected per leg lean mass (LLM). [(C)] time course of leg glucose uptake (data are means ± SEM). [(D)] leg glucose uptake during the steady-state period of the HE clamp. (E-H) Arteriovenous (a-v) difference in plasma glucose concentration and the femoral arterial blood flow corrected per leg lean mass (LLM). [(E) and (G)] time course of leg a-v glucose difference and leg blood flow (data are means ± SEM). [(F) and (H)] a-v glucose difference and leg blood flow during the steady-state period of the HE clamp. (I) Individual responses to the exercise training intervention, as calculated from the fold-difference between the trained and non-trained leg, in the relative contribution of glucose extraction and tissue perfusion to the changes in muscle glucose uptake. (J to L) Canonical insulin signaling as determined by phosphorylation of Akt2 on Ser473 and Thr308, and phosphorylation of the Akt substrates GSK3β on Ser9 and TBC1D4 on Thr 642 in whole-muscle lysates from skeletal muscle biopsy samples obtained before (Basal) and immediately after (Insulin) the hyperinsulinemic-euglycemic (HE) clamp. Levels of total Akt2, GSK3β, and TBC1D4 were comparable across legs and unaffected by insulin. Basal, n = 22; Insulin, n = 19 (10 in trained leg, 9 in non-trained leg). (M and N) mTORC1 and AMPK signaling as determined by phosphorylation of p70 S6 kinase (p70S6K) on Thr389 and AMPKα2 on Thr172 in whole-muscle lysates from skeletal muscle biopsy samples obtained before (Basal) and immediately after (Insulin) the HE clamp. Levels of total p70S6K were comparable across legs and unaffected by insulin. Basal, n = 22; Insulin, n = 19 (10 in trained leg, 9 in non-trained leg). Linear mixed models were used to estimate between-leg differences [(D)), (F), (H), (J), (K), (L), (M) and (N)]. Data are presented as observed individual values (with lines connecting the non-trained and trained leg from the same participant) and estimated means ± 95% confidence limits, unless otherwise stated. Individual datapoints are color-shaded to indicate muscle mtDNA heteroplasmy (light = low, dark = high). n = 22.n = 22, unless otherwise stated. *Different from Basal (*P* < 0.05). Illustrations in (A) were created with BioRender.com.

Exercise training improved muscle insulin sensitivity by ∼20%, as reflected by greater insulin-stimulated glucose uptake in the trained leg than in the contralateral non-trained leg (Cohen’s *d* = 0.86) (Fig. 6C-D). At the group level, this improvement was dissociated from significant changes in either arteriovenous glucose difference or leg blood flow (Fig. 6E-H). However, individual responses revealed marked heterogeneity in the relative contribution of these two components to the increase in muscle glucose uptake (Fig. 6I). Exercise-induced changes in glucose extraction and tissue perfusion tended to be inversely related across carriers (Pearson’s *r* = −0.59; *P* = 0.055), consistent with compensatory coupling of the two components. Leg-specific indirect calorimetry showed similar substrate oxidation rates in the trained and non-trained legs (Fig. S5A-C). By contrast, net plasma lactate release was higher in the trained leg (Fig. S5D-E), suggesting that, in the absence of a detectable increase in carbohydrate oxidation, part of the additional glucose taken up by the trained leg was directed toward non-oxidative disposal, including lactate production.

At the molecular level, insulin-stimulated phosphorylation of Akt, GSK3β, and TBC1D4 signaling nodes was similar between the trained and non-trained legs (Fig. 6J-L). Likewise, the exercise intervention did not alter insulin-stimulated mTORC1 or AMPK signaling (Fig. 6M and N).

Taken together, these data show that short-term exercise training ameliorates muscle IR in m.3243A>G carriers. This insulin-sensitizing effect was dissociated from enhanced signaling through the canonical Akt-TBC1D4 axis and from restoration of insulin-stimulated mTORC1 activity.

**Exercise training remodels the muscle mitochondrial proteomic and bioenergetic profile in m.3243A>G carriers**

PCA of muscle proteomes from the trained and non-trained legs showed an overall shift along PC1 (Fig. 7A), consistent with broad, albeit heterogenous, proteomic remodeling in response to the exercise intervention. Differential abundance analysis identified 108 proteins increased and 83 decreased in the trained relative to the non-trained leg (FDR < 0.05) (Fig. 7B). Mitochondrial proteins accounted for most exercise-induced increases (73/108), indicating robust mitochondrial remodeling in m.3243A>G carriers despite their inherent mitochondrial defect. AMPKγ2 (PRKAG2) was the most significantly increased protein, reversing the direction of its cross-sectional depletion. Other exercise-responsive proteins with metabolic relevance included BSG and CCDC47, involved in lactate flux and endoplasmic reticulum Ca^2+^ homeostasis, respectively, as well as the mitochondrial proteins TMEM70 and FAM210B (Fig. 7C). Among the proteins reduced by exercise was NQO2, which contributes to cellular redox regulation. INSR and GLUT4 abundance were unchanged, whereas hexokinase I and II showed nominal increases with exercise (unadjusted *P*<0.05) (Fig. 7D), in line with greater intramyocellular glucose-phosphorylating capacity.

**Figure 7.**
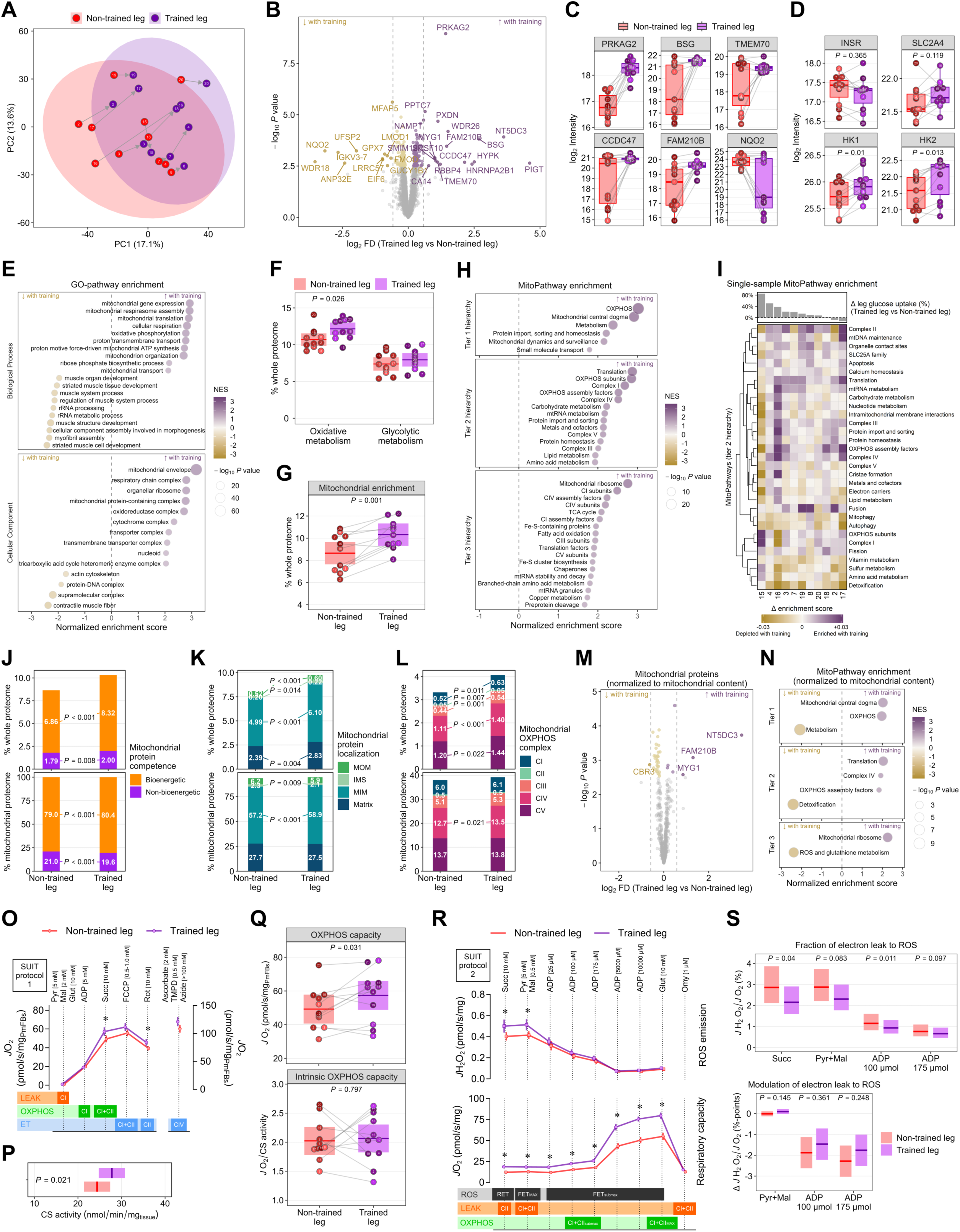
Exercise training remodels the muscle mitochondrial proteomic and bioenergetic profile in m.3243A>G carriers. (A) Principal component (PC) analysis of the muscle proteome in trained and non-trained legs of m.3243A>G carriers. (B) Volcano plot of differentially abundant muscle proteins in trained vs. non-trained legs. Downregulated and upregulated proteins in trained legs are marked in red and purple, respectively (FDR < 5%). (C) Boxplots of selected muscle proteins related to metabolism and differentially abundant in trained compared to non-trained legs (*P_adj_* < 0.05). (D) Abundance of insulin receptor (INSR), GLUT4 (SLC2A4), hexokinase I (HK1), and hexokinase II (HK2) in trained compared to non-trained legs (unadjusted *P* values are reported). (E) Bubble plots of most significantly depleted and enriched Gene Ontology (GO) Biological Process and Cellular Component terms in trained legs. Bubble plots are colored by normalized enrichment score, and size is based on *P*_adj_. (F) Proportion of oxidative and glycolytic metabolism proteins relative to the whole proteome. (G) Mitochondrial protein enrichment factor as determined by the summed mitochondrial protein abundance relative to the whole proteome. (H) Bubble plot showing significantly depleted and enriched mitochondrial pathways (MitoPathways from MitoCarta3.0) in trained legs. Bubble plots are colored by normalized enrichment score and size is based on *P_adj_*. (I) Hierarchically clustered heatmap showing matched trained vs. non-trained differences in MitoPathways (Tier 2) enrichment scores. The top bar plot indicates the corresponding difference in muscle insulin sensitivity (leg glucose uptake during the hyperinsulinemic-euglycemic clamp). (J) Proportion of bioenergetically (“*core bioenergetic*” and “*enabling bioenergetic*” annotated proteins) and non-bioenergetically competent mitochondrial protein abundance relative to the whole and mitochondrial proteome. (K) Proportion of compartment-specific mitochondrial protein abundance relative to the whole and mitochondrial proteome. MOM, mitochondrial outer membrane; IMS, intermembrane space; MIM, mitochondrial inner membrane. (L) Proportion of mitochondrial OXPHOS system proteins relative to the whole and mitochondrial proteome. (M) Volcano plot of differentially abundant mitochondrial proteins in trained vs. non-trained legs after normalization to mitochondrial protein content. (N) Significantly enriched and depleted MitoPathways in m.3243A>G carriers after normalization to mitochondrial protein content. (O) Substrate-inhibitor titration (SUIT) protocol 1 used to measure *ex vivo* mitochondrial respiratory capacity (O_2_ consumption rate, *J*O_2_) in permeabilized fiber bundles (PmFBs) (data in B are means ± SEM). (P) Citrate synthase (CS) activity in whole muscle homogenates (reflecting mitochondrial content in PmFBs) (Q) Mass-specific and intrinsic OXPHOS capacity in PmFBs as determined by *J*O_2_ in the presence of saturating concentrations of pyruvate, malate, glutamate, ADP, and succinate. (R) SUIT protocol 2 used to simultaneously measure *ex vivo* mitochondrial respiratory capacity and ROS emission (H_2_O_2_ emission rate, *J*H_2_O_2_) in PmFBs (data are means ± SEM). (S) Fraction of electron leak to ROS as determined by the ratio between mitochondrial H_2_O_2_ emission and O_2_ consumption rates, and modulation of electron leak to ROS by pyruvate/malate and ADP. Linear mixed models were used to estimate between-leg differences [(E), (F), (I), (J), (K), (O), (P), and (R)]. Data are presented as observed individual values (with lines connecting the non-trained and trained leg from the same participant) and estimated means ± 95% confidence limits. Individual datapoints are color-shaded to indicate muscle mtDNA heteroplasmy (light = low, dark = high). n = 22.

At the pathway level, GO-based enrichment analysis showed broad exercise-induced enrichment of mitochondrial processes and compartments together with depletion of myogenic and contractile pathways (Fig. 7E). This was accompanied by increased relative abundance of oxidative but not glycolytic metabolism proteins (Fig. 7F). Targeted mitochondrial analyses further revealed increased mitochondrial protein enrichment with exercise, together with broad enrichment of MitoPathways, including mitochondrial ribosome- and complex I-related signatures (Fig. 7G-H). Single sample-based enrichment analysis indicated that most carriers exhibited exercise-induced enrichment of OXPHOS-associated pathways, particularly OXPHOS subunits and complex I, together with depletion of detoxification-related pathways, although the magnitude of response varied across individuals (Fig. 7I).

To distinguish increased mitochondrial protein abundance from selective remodeling of the mitochondrial proteome, mitochondrial protein classes were quantified relative to either the whole proteome or the mitochondrial proteome (Fig. 7J-L). Exercise increased the abundance of all mitochondrial protein classes when expressed relative to the whole proteome. In contrast, when normalized to the mitochondrial proteome, enrichment persisted for bioenergetic and mitochondrial inner membrane proteins, whereas non-bioenergetic and intermembrane space proteins were reduced relative to the mitochondrial proteome, consistent with an exercise-induced shift toward a more bioenergetically competent mitochondrial proteome. After normalization to mitochondrial protein content, only 7 mitochondrial proteins remained increased, whereas 31 proteins appeared reduced with exercise (FDR < 0.05) (Fig. 7M). MitoPathways enrichment analysis on the normalized mitochondrial proteome retained enrichment of translation- and OXPHOS-related pathways together with depletion of detoxification-related programs (Fig. 7N), supporting selective mitochondrial proteome remodeling beyond an increase in mitochondrial content *per se*.

At the functional level, muscle mitochondrial bioenergetic analyses showed that exercise increased mass-specific, but not intrinsic, OXPHOS capacity, indicating that this functional improvement stemmed from increased mitochondrial content (Fig. 7O-Q). These changes were accompanied by a lower fraction of electron leak to ROS (Fig 7R-S), which however was not reflected in alterations in mitochondrial redox state (Fig S6). Collectively, these findings are consistent with exercise acting as a potent modifier of mitochondrial abnormalities in m.3243A>G muscle and provide first-in-human mechanistic evidence suggesting mitochondrial remodeling as a key contributor to improved muscle insulin sensitivity.

### Exercise reverses interconnected mitochondrial and AMPKγ2 defects associated with muscle insulin action

To systematically test whether exercise reverses the proteomic alterations identified in the cross-sectional cohort, we computed a rescue statistic quantifying directional opposition between exercise-induced changes and m.3243A>G-related defects at three hierarchical levels: GO Biological Process and Cellular Component pathways, MitoPathways, and individual proteins. At the biological process level, pathways most affected by m.3243A>G shifted in the opposite direction with exercise, indicating strong directional reversal (Fig. 8A). Cellular respiration and oxidative metabolism pathways were among the most depleted in m.3243A>G and the most strongly rescued by exercise. A concordant pattern emerged at the cellular component level, with mitochondrial compartments among the most prominently rescued by exercise (Fig. S7). Within the mitochondrial proteome, most MitoPathways displayed a consistent pattern of m.3243A>G-related depletion and exercise-induced enrichment, with OXPHOS, TCA cycle, and mitochondrial translation pathways among the most strongly rescued (Fig. 8B). At the individual protein level, m.3243A>G-related defects and exercise-induced effects were positively correlated, with PRKAG2 showing the strongest combined signature of m.3243A>G-linked downregulation and exercise-induced upregulation (Fig. 8C).

**Figure 8.**
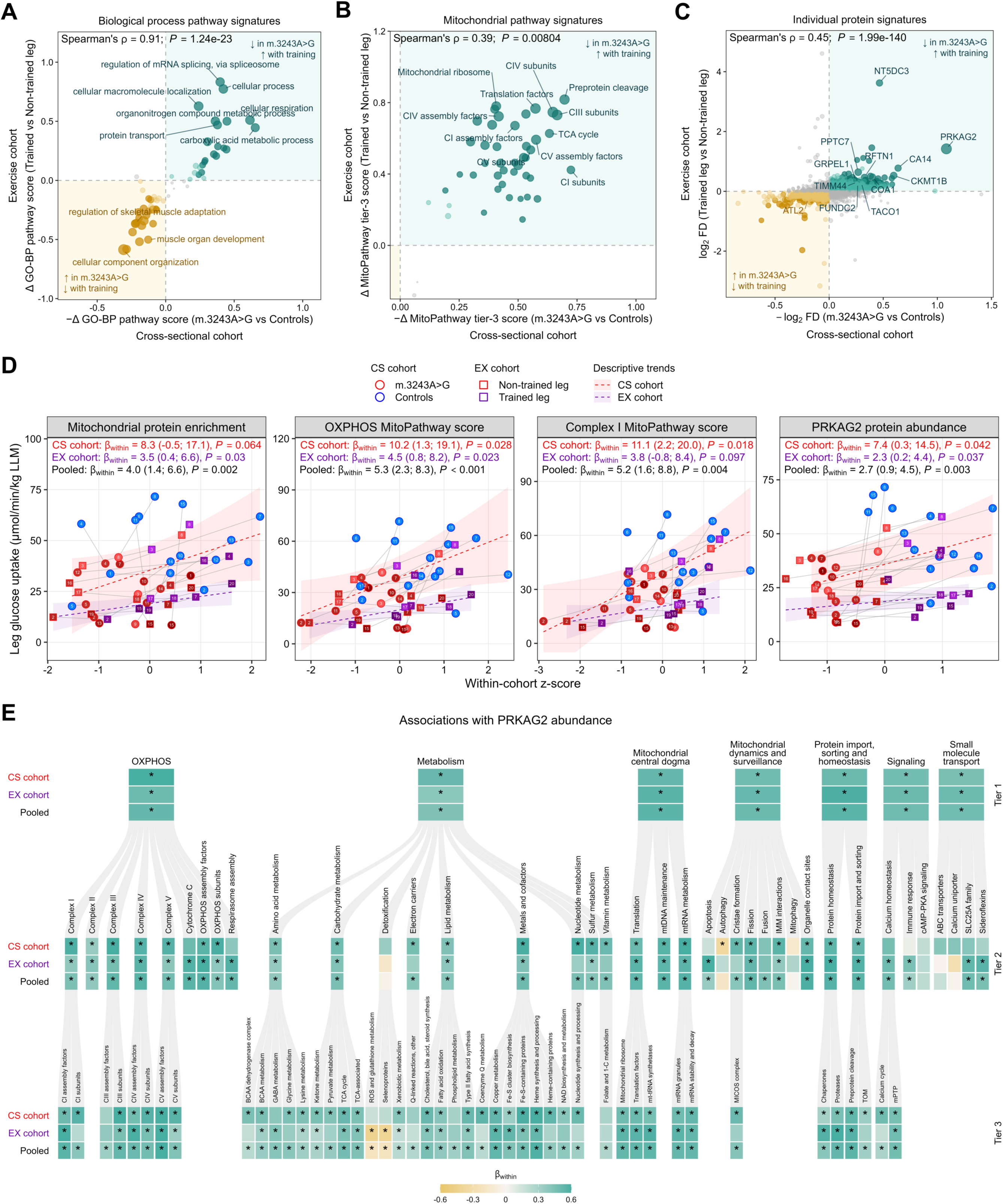
Exercise reverses interconnected mitochondrial and AMPKγ2 defects associated with muscle insulin action. (A-C) Cross-study integration of m.3243A>G variant defects (cross-sectional cohort, x-axis) and exercise training effects (exercise cohort, y-axis) at three hierarchical levels. Each scatter plot displays the negated cross-sectional effect (−Δ pathway or protein score; m.3243A>G vs Controls) against the exercise effect (Δ pathway or protein score; trained leg vs non-trained leg), such that features in the upper-right quadrant were suppressed in carriers and rescued by exercise, and features in the lower-left quadrant were elevated in carriers and reduced by exercise. Point size reflects the rescue significance (−log_10_ *P* rescue). Spearman’s ρ and *P* values are shown. [(A)] GO Biological Process pathway signatures (mean-z scores). [(B)] MitoCarta tier-3 MitoPathway signatures (mean-z scores). [(C)] Individual protein signatures (log_2_ fold differences). Teal points: pathways/proteins downregulated in carriers and upregulated by exercise; dark teal: FDR<0.05. Gold points: pathways/proteins upregulated in carriers and downregulated by exercise; dark gold: FDR<0.05. (D) Associations between pre-defined molecular signatures and insulin-stimulated leg glucose uptake in the cross-sectional (CS) and exercise (EX) cohorts. Analyses were conducted within-clusters, defined as matched pairs (m.3243A>G vs Controls) in the CS cohort or as participants (trained vs non-trained leg) in the EX cohort; the reported β_within_ coefficients reflect covariation within these clusters. For each molecular signature, protein abundance was standardized within cohort (z-scored) and a fixed-effects model including cluster as a covariate was fitted separately per study. Cohort-specific β_within_ values represent the change in leg glucose uptake per 1-SD within-cohort increase in the predictor within a cluster, with 95% CI. Pooled β_within_ values were obtained by inverse-variance fixed-effect meta-analysis of the two study-specific estimates. Dashed lines and shaded bands represent cohort-specific marginal trend lines and 95% CI derived from the corresponding fixed-effects models. Thin grey lines connect observations within clusters. (E) Hierarchical heatmap of within-cluster associations between PRKAG2 protein abundance and MitoCarta tier-2 MitoPathway scores, estimated using the same framework as in (D). *FDR<0.05.

We next interrogated whether the molecular signatures most affected by m.3243A>G and most strongly rescued by exercise were associated with muscle insulin sensitivity. To this end, we estimated associations with insulin-stimulated leg glucose uptake within matched carrier-control pairs in the cross-sectional cohort and within participants across legs in the exercise cohort, and then pooled the results across cohorts. Greater mitochondrial protein enrichment, OXPHOS and complex I pathway scores, and PRKAG2 protein abundance were each positively associated with greater muscle insulin sensitivity (Fig. 8D). By contrast, no significant associations were observed with insulin-stimulated mTORC1 or AMPK signaling readouts (Fig. S7).

We then examined the relationship between PRKAG2 and mitochondrial signatures. PRKAG2 abundance was positively associated with the large majority of mitochondrial pathways, including all OXPHOS, carbohydrate metabolism, and mitochondrial translation pathways (Fig. 8E). Notable exceptions included autophagy, mitophagy, and detoxification pathways, suggesting that PRKAG2 specifically tracks with mitochondrial bioenergetic rather than quality-control programs. This pattern was consistent across cohorts and across pathway hierarchical levels, indicating robust coordinate regulation of PRKAG2 and the mitochondrial bioenergetic program, regardless of whether the underlying variation arose from m.3243A>G or from exercise. Together, these analyses show that exercise partly reverses m.3243A>G-related proteomic defects linked to muscle IR and identify PRKAG2 as a prominent molecular correlate of both mitochondrial remodeling and insulin sensitivity in human skeletal muscle.

## DISCUSSION

To probe the causal contribution of mitochondrial dysfunction to human insulin resistance (IR) and characterize the underlying molecular mechanisms, we studied individuals carrying the rare m.3243A>G mtDNA variant, an inherited cause of mitochondrial dysfunction associated with a high prevalence of diabetes. Deep *in vivo* cross-sectional phenotyping showed selective skeletal muscle IR with preserved liver and adipose tissue insulin sensitivity in m.3243A>G carriers. At the molecular level, this muscle insulin-resistant phenotype was characterized by preserved insulin signaling through the Akt-TBC1D4 axis but impaired insulin-stimulated mTORC1 activation. Skeletal muscle proteomic profiling revealed broad depletion of the mitochondrial proteome, disproportionately affecting OXPHOS- and complex I-related pathways, alongside marked reduction of AMPKγ2 abundance. Functionally, muscle mitochondrial bioenergetic analyses demonstrated reduced OXPHOS capacity largely attributable to lower mitochondrial content, while also uncovering complex I-specific impairments independent of mitochondrial content. An exercise-based mechanistic intervention in m.3243A>G carriers showed that exercise improved muscle insulin sensitivity alongside partial restoration of mitochondrial protein abundance, OXPHOS- and complex I-specific signatures, and AMPKγ2 abundance, without parallel enhancement of insulin-stimulated Akt-TBC1D4 or mTORC1 signaling. An analytical framework integrating cross-sectional phenotypic features and exercise-induced remodeling signatures confirmed that molecular alterations characteristic of m.3243A>G muscle were rescued by exercise, with mitochondrial proteome enrichment, OXPHOS and complex I signatures, and AMPKγ2 abundance each positively associated with muscle insulin sensitivity.

Our comprehensive multi-tissue *in vivo* characterization of insulin action not only corroborate previous evidence of muscle insulin resistance in m.3243A>G carriers^17^, but also demonstrate that this impairment is confined to skeletal muscle, with no apparent defects in liver or adipose tissue insulin action. This selective muscle insulin-resistant phenotype, together with our and prior observations of systemic IR in m.3243A>G carriers^17,18,22,49,50^, indicates that muscle IR is a primary driver of the reduced insulin-stimulated whole-body glucose disposal in this population. Importantly, muscle IR in m.3243A>G carriers was attributable to both reduced muscle glucose extraction and diminished insulin-stimulated blood flow, the latter indicating impaired endothelial-mediated vasodilation and thus aligning with evidence implicating endothelial dysfunction as a critical contributor to muscle IR^83–85^. This finding raises the possibility that inherent defects in endothelial mitochondria may contribute to the observed insulin-resistant phenotype, extending the detrimental impact of the m.3243A>G variant to vascular compartments upstream of the myocyte. This notion is further supported by the observation that the exercise-induced amelioration of muscle IR in m.3243A>G carriers stem from heterogenous improvements in either muscle glucose extraction or tissue perfusion. Together, these findings establish individuals with the rare m.3243A>G variant as a human genetic model for dissecting mitochondrial mechanisms driving muscle IR.

To define the molecular basis of mitochondria-linked IR, we combined in-depth proteomic profiling with bioenergetic analyses of skeletal muscle biopsies from m.3243A>G carriers. This addressed an important translational gap, as current evidence for m.3243A>G-associated mitochondrial defects derives predominantly from cultured cell models^46–48,54–59^, which do not fully recapitulate the physiological architecture, metabolic environment, and regulatory cues governing muscle mitochondrial biology *in vivo*.

The observed depletion of the muscle mitochondrial proteome in m.3243A>G carriers is consistent with the role of *MT-TL1* in mitochondrial protein synthesis. This finding indicates that reduced mitochondrial content is a characteristic feature of the muscle insulin-resistant phenotype associated with m.3243A>G, and directly aligns with evidence linking lower mitochondrial content to whole-body^35^ and muscle^25,60^ IR. Our pathway-specific analysis of the mitochondrial proteome reveals that the m.3243A>G variant does not uniformly reduce mitochondrial protein abundance but instead selectively reshapes mitochondrial proteomic composition, with a disproportionate depletion of bioenergetic programs and enrichment or preservation of non-bioenergetic pathways. This divergent regulation suggests a functional remodeling of mitochondria toward non-bioenergetic processes, potentially reflecting compensatory responses to impaired OXPHOS capacity. Importantly, these mitochondrial proteomic defects were partially reversible, with exercise increasing mitochondrial protein enrichment, rescuing mitochondrial pathway signatures, and shifting the proteome toward a more bioenergetically competent composition. The correspondence between the magnitude of m.3243A>G-related defects and exercise-mediated rescue across the muscle proteome further supports the functional relevance of these mitochondrial signatures to the insulin-resistant phenotype.

At the functional level, the finding that *ex vivo* mitochondrial respiratory capacity, specifically maximal OXPHOS and electron transport capacity in permeabilized muscle fibers, was reduced in carriers aligns with prior observations in fibroblasts^46^ and supports the presence of OXPHOS defects arising from the m.3243A>G variant. However, these defects were no longer apparent when accounting for the reduced mitochondrial content characterizing m.3243A>G muscle, indicating that mitochondrial respiratory function per mitochondrion was preserved. In the context of mitochondria-linked IR, this finding is consistent with prior evidence indicating preserved intrinsic respiratory capacity in skeletal muscle^23,61–65^, liver^66^, and adipose tissue^67^ of insulin-resistant individuals. Similarly, we observed that mitochondrial ROS emission capacity was reduced in carriers but remained proportional to their lower mitochondrial respiratory capacity, indicating no disproportionate electron leak to ROS. This finding, together with the absence of alterations in mitochondrial redox state (PRDX3 dimerization), suggests that, unlike lipid-induced IR where mitochondrial oxidative stress plays a causal role^29^, perturbations in mitochondrial redox homeostasis have a limited role in muscle IR associated with chronic mitochondrial defects. Consistent with these observations, exercise increased OXPHOS capacity in m.3243A>G carriers, but this effect was attributable to increased mitochondrial content rather than improved intrinsic respiratory function. Notably, however, exercise also reduced the fractional electron leak to ROS, indicating that the increase in mitochondrial content was accompanied by qualitative improvements in electron transport chain coupling. Together with the absence of changes in mitochondrial redox state, these findings suggest that exercise enhances mitochondrial bioenergetic capacity in m.3243A>G carriers predominantly through quantitative expansion and compositional remodeling of the mitochondrial proteome, rather than through amelioration of intrinsic respiratory function or redox state.

Taken together, our combined *in vivo* and *ex vivo* muscle phenotyping of m.3243A>G carriers provides human evidence that primary quantitative and bioenergetic mitochondrial defects contribute to muscle IR. The partial exercise-induced rescue of these defects alongside improved insulin sensitivity further challenges the concept, derived largely from pre-clinical and short-term clinical studies, that muscle mitochondrial dysfunction arises as a downstream consequence of IR^68,69^.

By integrating the proteomic and bioenergetic data, we identified a defective mitochondrial complex I signature in m.3243A>G carriers. Because m.3243A>G is a mtDNA variant, a predominant complex I defect might be expected, as 7 out of the 13 mtDNA-encoded OXPHOS proteins belong to complex I. However, the finding that both mtDNA- and nDNA-encoded OXPHOS proteins were reduced in carriers points to a preferential vulnerability of complex I rather than isolated loss of mtDNA gene products. These findings highlight a potential role of mitochondrial complex I in modulating muscle insulin action in humans, in line with a human study showing a positive correlation between complex I proteins and insulin sensitivity^70^ and with preclinical evidence that muscle-specific complex I deficiency impairs insulin signaling and glucose homeostasis^71,72^. Notably, the complex I defects observed here were accompanied by enrichment of myogenic and muscle-specific pathways, mirroring the enhanced myogenesis reported in complex I-deficient myotubes^72^, and thereby suggesting a conserved compensatory myogenic response to complex I dysfunction. The exercise intervention provides convergent support for the relevance of complex I to muscle biology. At the proteomic level, exercise enriched complex I pathways while suppressing myogenic and contractile programs. Importantly, the cross-study integration framework showed that complex I pathway enrichment was positively associated with muscle insulin sensitivity, providing quantitative support for a mechanistic link between mitochondrial complex I and muscle insulin action.

At the molecular level, mitochondrial dysfunction has traditionally been proposed to promote IR through impaired lipid oxidation, intramyocellular lipid accumulation, and consequent disruption of proximal insulin signaling^73^. However, this model does not fully explain the phenotype observed here. The preservation of insulin-stimulated Akt, GSK3, and TBC1D4 signaling in m.3243A>G carriers, together with the absence of exercise-induced enhancement in these nodes despite improved muscle insulin sensitivity, suggests that chronic mitochondrial dysfunction impairs insulin action through mechanisms beyond the canonical Akt-TBC1D4 axis regulating GLUT4 translocation. Consistent with this, carriers exhibited blunted insulin-stimulated mTORC1 activation, aligning with emerging evidence that mTORC1-linked signaling nodes, rather than canonical Akt substrates, are selectively impaired in insulin-resistant muscle^35,37^. This defective mTORC1 response may represent a mitochondrial dysfunction-associated feature of insulin-resistant skeletal muscle, as electron transport chain defects can activate branches of the integrated stress response that induce ATF4-dependent repression of mTORC1^74^. However, because exercise did not rescue insulin-stimulated mTORC1 activation, and mTORC1 activity was not associated with muscle insulin sensitivity in the present study, blunted mTORC1 activation is more likely an accompanying signature than a mediator of mitochondrial dysfunction-associated IR.

A second distinctive molecular feature of m.3243A>G muscle was marked downregulation of the regulatory γ2 subunit of AMPK (PRKAG2). This defect was reversed by exercise, and AMPKγ2 abundance correlated positively with both muscle insulin sensitivity and mitochondrial proteome signatures, linking AMPKγ2 to the broader mitochondrial phenotype and raising the possibility that its downregulation forms part of a mito-nuclear retrograde response to chronic mitochondrial stress^75^. Because the γ2 subunit contributes to AMPK complex composition and regulation of activation-loop phosphorylation^76,77^, reduced AMPKγ2 abundance could plausibly influence AMPK signaling output. However, any link between AMPKγ2 and insulin action appears more consistent with chronic regulation of muscle metabolism than with altered acute AMPK activation. In support, human genetic data associate *PRKAG2* polymorphism with IR^78^, whereas γ2 appears dispensable for acute pharmacological AMPK-stimulated muscle glucose uptake in rodents^79^. Consistent with this distinction, insulin-stimulated AMPKα2^Thr172^ phosphorylation was unaltered in carriers and unaffected by exercise, indicating that AMPKγ2-associated differences were not mirrored by alterations at this canonical AMPK activation site. Together, these findings identify AMPKγ2 as a candidate muscle-intrinsic node linking chronic mitochondrial dysfunction to IR.

Finally, the mitochondria-hexokinase II (HKII) axis may represent a complementary mechanism by which mitochondrial defects impair insulin-stimulated glucose uptake despite preserved Akt-TBC1D4 signaling. Mitochondria-bound HKII couples mitochondrial ATP production to glucose phosphorylation, thereby enhancing HKII catalytic activity^80^. Under insulin-stimulated conditions, constrained HKII capacity would be expected to increase intracellular free glucose, dissipate the transmembrane glucose gradient, and ultimately limit glucose uptake despite preserved insulin signaling^81,82^. Within this framework, reduced mitochondrial content and OXPHOS defects in m.3243A>G muscle may constrain HKII activity and contribute to impaired insulin-stimulated glucose uptake. Conversely, the observed exercise-induced mitochondrial remodeling, together with nominal increases in HKII protein abundance, is compatible with improved capacity of the mitochondria-HKII axis.

Collectively, these findings suggest that blunted insulin-stimulated mTORC1 activation, AMPKγ2 depletion, and a potential disruption of the mitochondria-HKII axis represent convergent features of a shared mitochondrial stress phenotype, thus supporting a model in which mitochondrial dysfunction influences muscle insulin action through parallel mechanisms that extend beyond the canonical Akt-TBC1D4 pathway.

### Limitations

This study is characterized by a relatively small sample size, reflecting both the rarity of the m.3243A>G variant and the experimental physiological design, which prioritized deep *in vivo* phenotyping and mechanistic insight through highly invasive clinical procedures and gold-standard assessments. While the study was adequately powered for its primary outcome (insulin sensitivity), statistical power for other outcomes may have been more limited. To reduce potential sources of variability, carriers and controls were rigorously matched for major biological confounders, including age, sex, body composition, and objectively measured habitual physical activity. Furthermore, m.3243A>G carriers with severe muscle involvement limiting ambulation or with comorbidities requiring pharmacological treatment were excluded a priori, ensuring appropriate matching with healthy controls and minimizing lifestyle- and pharmacotherapy-related confounding effects. This conservative approach reinforces the validity of the present findings, as marked physiological alterations were observed even in a mildly affected cohort of m.3243A>G carriers.

While this study leverages individuals with primary mitochondrial disorders as a human genetic model to interrogate the causal contribution of mitochondrial dysfunction to IR, diabetes is not uniformly prevalent across mitochondrial diseases. This variability reflects the phenotypic heterogeneity of mitochondrial diseases and indicates that only specific mitochondrial defects may drive IR and thereby contribute to diabetes pathophysiology. In this regard, while we identified a link between complex I-related defects and IR in m.3243A>G carriers, diabetes is not particularly common among individuals harboring mtDNA variants that affect complex I-specific subunits^15^. By contrast, pathogenic mitochondrial tRNA variants (including m.3243A>G) or single mtDNA deletions, which impair global mitochondrial protein synthesis, are more frequently associated with diabetes^15,22,83–86^. These observations indicate that broad mitochondrial perturbations, in addition to complex I-specific impairments, may be required to elicit detrimental effects on insulin action. Finally, given the multifaceted nature of mitochondrial biology, our phenotyping approach could not capture its full breadth. Future studies should therefore extend integrative physiological and molecular profiling to additional dimensions of mitochondrial biology and to other diabetogenic mtDNA variants to determine whether they exhibit mitochondrial signatures similar to those observed in m.3243A>G carriers, thereby refining the concept of a mitochondrial dysfunction fingerprint underlying IR.

### Conclusion and perspectives

This proof-of-concept physiological study leverages m.3243A>G carriers as a human genetic model of primary, chronic mitochondrial dysfunction to delineate mitochondrial mechanisms implicated in human IR. By integrating cross-sectional phenotyping with an exercise-based mechanistic intervention, we demonstrate that mitochondrial proteome depletion, OXPHOS- and complex I-specific defects, and reduced AMPKγ2 abundance are defining features of skeletal muscle IR in m.3243A>G carriers, and that these defects are partially rescued by exercise alongside improved muscle insulin sensitivity. This integrated defect-rescue framework strengthens the causal link between mitochondrial dysfunction and impaired insulin action in humans and establishes an exercise-modifiable mitochondrial basis for muscle IR that occurs independently of detectable defects in canonical Akt-TBC1D4 signaling.

Clinically, these findings position mitochondrial dysfunction as a tractable upstream determinant of impaired muscle insulin sensitivity. Although they cannot be readily generalized to common insulin-resistant states, the frequent association between mitochondrial dysfunction, IR, and T2DM supports mitochondria-targeted strategies as a rational therapeutic avenue beyond primary mitochondrial disease. PPAR agonists such as thiazolidinediones stimulate mitochondrial biogenesis and oxidative metabolism and remain among the most effective insulin-sensitizing agents^87^, but their systemic benefits are largely adipose tissue-mediated^88^, leaving unresolved whether muscle-targeted PPAR modulation provides additional benefit^89^. Emerging agents targeting mitochondrial biogenesis and/or quality control, including NAD⁺ precursors, (–)-epicatechin, and urolithin A, show skeletal muscle target engagement in humans^90–93^, yet evidence for corresponding improvements in insulin sensitivity remains limited. Finally, resolving whether AMPKγ2 is a mediator, marker, or adaptive consequence of mitochondrial dysfunction-related IR may clarify whether this regulatory AMPK subunit represents a tractable node for jointly restoring mitochondrial function and insulin action in human skeletal muscle.

## Data Availability

All data produced in the present study are available upon reasonable request to the authors.

## ACKNOWLEDGEMENTS

We thank all the study participants. We are grateful to Thomas Krag (Copenhagen Neuromuscular Center, Department of Neurology, Copenhagen University Hospital–Rigshospitalet) for excellent technical support and to Mikkel Taudorf and colleagues (Department of Interventional Radiology, Copenhagen University Hospital–Rigshospitalet) for assistance and supervision during the clinical experimental procedures. Proteomic sample preparation, LC-MS measurements, and data analysis were performed by the Proteomics Research Infrastructure at the University of Copenhagen, supported by the Novo Nordisk Foundation (grant agreement number NNF19SA0059305). The proteomics data have been deposited to the ProteomeXchange Consortium via the PRIDE partner repository with the dataset identifier PXD073394 (cross-sectional study) and PXD077576 (exercise intervention study). During the preparation of this work the authors used generative AI and AI-assisted technologies to edit R scripts and improve manuscript text for grammar and style. After using this tool, the authors reviewed and edited the content as needed and take full responsibility for the content of the published article.

## AUTHOR CONTRIBUTIONS

Conceptualization: J.V., and M.F.;

Data curation: T.L.N., and M.F.;

Formal analysis: T.L.N., M.Da., and M.F.;

Funding Acquisition: L.S., J.V., and M.F.;

Investigation (clinical): T.L.N., M.Da., and M.F.;

Investigation (biological sample analyses): T.L.N., M.Da., N.R.A, M.Du., G.v.H., L.S., and M.F.;

Methodology: M.M., S.L., S.S.T, J.V., and M.F.;

Project administration: T.L.N., and M.F.;

Resources: A.L.F., G.v.H, L.S., S.L., S.S.T., and J.V.;

Software: M.F.;

Supervision: S.L., J.V., and M.F.;

Validation: T.L.N., and M.F.; Visualization: T.L.N., and M.F.;

Writing – Original Draft: T.L.N, and M.F.;

Writing – Review and Editing: T.L.N., M.Da., N.R.A., M.Du., A.L.F., G.v.H., L.S., M.M., S.L., S.S.T., J.V., and M.F.

All authors contributed to data interpretation, critically revised the manuscript for important intellectual content and approved the final version of the manuscript.

## METHODS

### Study design and ethical statement

In a cross-sectional study design, fifteen adults carrying the m.3243A>G variant and fifteen healthy persons attended three study visits consisting of one screening visit and two experimental trial days. To control for potential confounding effects of aging, sex hormones, adiposity, and lifestyle factors, m.3243A>G carriers and controls were individually matched for age, sex, body mass index (BMI), and objectively measured physical activity (Table S1). Matching for BMI and physical activity accounted for prior reports of reduced body weight and lower habitual activity in individuals with mitochondrial disorders^94,95^.

In a separate within-participant longitudinal intervention study design, eleven adults carrying the m.3243A>G variant attended a screening visit and underwent an exercise training period followed by one post-intervention study visit. Participants were instructed to maintain their habitual diet and physical activity throughout the study period. The studies were approved by the Research Ethics Committee of the Capital Region of Denmark (H-23003677) and conducted in accordance with the Declaration of Helsinki. All participants received written and verbal information about the study and provided written informed consent prior to any screening procedures. The studies were registered at ClinicalTrials.gov (identifier: NCT06080581 and NCT06080594).

### Study participants

#### Cross-sectional cohort

Individuals carrying the m.3243A>G variant were recruited from the Copenhagen Neuromuscular Center, a national referral center for mitochondrial disorders, and the Department of Clinical Genetics at the Copenhagen University Hospital-Rigshospitalet. A total of 102 carriers were identified and invited, 15 of whom were included in the study (Fig. S8). Inclusion criteria were: i) confirmed heteroplasmy for m.3243A>G mtDNA previously identified on genetic testing, and ii) age ≥18 years. Exclusion criteria were: i) use of antiarrhythmic medications or a diagnosis of severe heart disease (symptomatic ischemic heart disease, heart failure, atrial flutter or atrial fibrillation with EHCA-score >1, moderate to severe aorta stenosis, symptomatic anemia), ii) any endocrine disorder other than diabetes, iii) pregnancy, or iv) other conditions or treatment with medications that could affect study outcomes. Heteroplasmy (i.e. the ratio of mutated-to-wild type mtDNA) level in blood and muscle tissue was known *a priori* for 7 (blood) and 2 (muscle) carriers.

Healthy controls were recruited through a call on online recruitment platforms. Exclusion criteria were: i) age <18 years, ii) current or regular use of antidiabetic medications or other medications affecting study outcomes, iii) prior medical history of heart, lung, kidney, liver or endocrine conditions affecting study outcomes, iv) daily use of tobacco products, v) excessive alcohol consumption (above 20 drinks per week), vi) pregnancy, or vii) major lifestyle changes during the three months prior to the study period.

For females, menopausal status was determined based on frequency and regularity of menstrual cycle, usage of intrauterine devices preventing menstruation, and climacteric symptoms.

#### Exercise intervention cohort

Following the cross-sectional study, six carriers were enrolled in the longitudinal interventional study. Five additional carriers were recruited from the Copenhagen Neuromuscular Center and the Department of Clinical Genetics at the Odense University Hospital using the same inclusion and exclusion criteria described above.

### Screening visit

Carriers who accepted the invitation to participate in the study underwent a medical interview and a screening examination with a physician, including lung and heart auscultation, resting electrocardiography (ECG), resting blood pressure measurements, fasting blood samples (≥8 h fast), and a whole-body dual-energy X-ray absorptiometry (DXA)-scanning to assess body composition. Thereafter, carriers completed an incremental test to volitional exhaustion on a mechanically braked cycle-ergometer (Lode, Groningen, The Netherlands) to determine peak oxygen consumption (V̇O_2_peak). The protocol consisted of a 4-min warm-up at 40-80 W followed by an incremental phase with increments of 10-20 W/min according to individual exercise tolerance and fitness, aiming to achieve exhaustion within ∼10-15 min of the incremental phase. Pulmonary gas exchanges were measured breath-by-breath using an online gas analyser (CPET Quark-system, Cosmed, Italy). Finally, carriers enrolled in the cross-sectional study were provided with triaxial wrist-worn accelerometers (GENEActiv, Activinsights Ltd., Cambridgeshire, UK) to objectively measure habitual physical activity over 7 days of an ordinary week. Raw accelerometry data were processed using the *GGIR* package^96^. Sedentary time and time spent on light-, moderate-, and vigorous-intensity physical activity was calculated using previously validated activity thresholds^97^.

For candidate healthy controls in the cross-sectional study, 171 volunteers were pre-screened based on sex, age, and BMI. From this group, 59 individuals met the initial inclusion criteria and were provided with wrist-worn accelerometers to objectively determine habitual physical activity, as described for the carriers. Final matching criteria included an age difference within ±10 years and a moderate-to-vigorous physical activity level within ±10% (minutes/day) of the corresponding carrier. Healthy control candidates who met these matching criteria were subsequently invited to the screening visit. During the medical interview, healthy controls were additionally screened for diabetes family history and for clinical signs of the m.3243A>G variant, such as hearing impairment or MELAS features.

### Study visits

#### Cross-sectional study

Participants attended two study visits interspersed by at least three days (mean: 21 days). Study visits consisted of either an oral glucose tolerance test (OGTT) or a hyperinsulinemic-euglycemic clamp combined with skeletal muscle biopsies. Participants were instructed to refrain from caffeine, alcohol, and exercise for 48 hours before each visit and reported to the laboratory following an overnight fast (≥8 h). Two carriers undergoing pharmacological treatment for diabetes discontinued oral glucose-lowering medications for at least 4 days and withheld fast-acting insulin for at least 4 h prior to each study visit.

#### Exercise intervention study

Participants attended one experimental trial day consisting of a hyperinsulinemic-euglycemic clamp combined with skeletal muscle biopsies. Participants refrained from caffeine, alcohol, and exercise for 48 hours before the study visit and reported to the laboratory following an overnight fast (≥8 h). Four carriers undergoing pharmacological treatment for diabetes discontinued oral glucose-lowering medications for at least 4 days and withheld fast-acting insulin for at least 4 h and long-acting insulin for at least 24h prior to the study visit.

### Longitudinal exercise intervention

The longitudinal intervention consisted of a short-term single-leg cycling high-intensity interval training (HIIT) protocol comprising eight sessions separated by 2-3 days. Participants were provided with mechanically-braked ergometer bikes (Monark 928E, Vansbro, Sweden) modified for single-leg cycling, i.e. a 10-kg counterweight was attached to the unoccupied crank arm to assist the upstroke of the active leg and maintain crank inertia, while the contralateral leg remained inactive. Upon delivery, ergometers were calibrated according to manufacturer’s instructions and participants were familiarized with the setup. Thereafter, participants completed a single-leg graded exercise test to determine incremental peak power output (iPPO). The test started at 30-40 W with increments of 5-10 W/min depending on the participant’s fitness level. Single-leg iPPO and maximal heart rate achieved during the test were used to individualize exercise intensity during the intervention. Each training session consisted of a 5-min warm-up at 20% iPPO, followed by 5-min high-intensity intervals at 65% iPPO interspersed with 2.5-min active recovery at 20% iPPO. The number of high-intensity intervals increased progressively from two to four across the intervention period (Fig. 6A). Training sessions were supervised online, and heart rate was continuously monitored (Vivosmart 4, Garmin, Kansas, USA). Workload was adjusted when necessary based on participant feedback and heart rate responses to maintain the intended exercise intensity while ensuring completion of each session. Participants were instructed to maintain their habitual lifestyle and daily physical activity throughout the study.

### Oral glucose tolerance test

Upon arrival, participants rested in the supine position for 30 min before placement of a catheter in an antecubital vein. After baseline blood sampling, participants ingested 75 g glucose dissolved in 300 mL of water. Participants remained seated throughout the test, and venous blood samples were collected at 15, 30, 45, 60, 75, 90, 120, and 180 min.

### Hyperinsulinemic-euglycemic clamp

Catheters (Micropuncture Introducer Set, Cook Medical, Bloomington, Indiana, USA) were inserted into the femoral artery and vein of the right leg under local anesthesia (10% Lidocaine, Amgros, Denmark) for arterial and venous blood sampling. Correct positioning was verified by ultrasound imaging (Sonosite PX, Fujifilm, USA). A separate catheter was placed in an antecubital vein for infusions. Thereafter, a primed (2.6 mg/kg) constant infusion of [6,6-^2^H_2_]glucose tracer (0.044 mg/kg/min; Cambridge Isotope Laboratories, Massachusetts, USA) was initiated and continued for 2 h before starting the hyperinsulinemic-euglycemic clamp. The clamp began with a bolus of insulin (9 mU/kg; Actrapid, Novo Nordisk, Denmark) followed by a constant insulin infusion for 150 min (1.42 mU/kg/min). To prevent non-specific insulin binding to the infusion lines and syringes, insulin was diluted in saline containing 10% of the participant’s own blood. During the clamp, glucose was infused from a 10% glucose solution (Fresenius-Kabi) enriched with [6,6-^2^H_2_]glucose. Arterial plasma glucose was measured every 5 min during the first 30 min of the clamp and every 7.5 min thereafter, with glucose infusion rates adjusted to maintain euglycemia (5.5 mM). Femoral artery blood flow measurements and paired femoral arterial-venous blood samples were obtained before tracer infusion (−140 min), immediately before initiation of the clamp (−5 min), and at 30, 60, 90, 105, 120, 135, and 150 min during the clamp.

For the exercise intervention study, the hyperinsulinemic-euglycemic clamp protocol was similar except for that i) catheters were inserted into the femoral artery of a randomized leg and into femoral veins of both legs, and ii) [6,6-^2^H_2_]glucose tracer was not infused.

### Femoral artery blood flow

Femoral artery blood flow was measured by Doppler ultrasound (Sonosite PX, Fujifilm, USA) using a linear probe operating at an imaging frequency of 8.0 MHz and Doppler frequency of 5.3 MHz. At each time-point, three consecutive 10-s recordings were acquired, and the mean of the three replicate measurements was used for analysis. For the longitudinal intervention study, femoral artery blood flow was measured bilaterally at each time-point.

### Leg lean mass

Lean mass of the experimental leg (defined as fat-free mass minus bone mineral mass) was assessed by DXA (Lunar Prodigy Advance, GE Healthcare, USA) by defining the leg region as the area extending from the inferior border of the ischial tuberosity to the distal tip of the toes.

### Blood and plasma analyses

During the OGTT trial day, venous blood samples were drawn in BD Emerald syringes (BD, New Jersey, USA) and transferred to spray-coated EDTA Tubes (BD Vacutainer, BD). Samples were centrifuged at 10,000 *g* for 10 min at 4° C, and the resulting plasma was aliquoted and stored at −80 °C for subsequent analyses of glucose, insulin, C-peptide, and free fatty acids (FFA). Plasma glucose was quantified using the GLUC3-kit on a Cobas 503 system (Roche Diagnostics, Mannheim, Germany). Plasma insulin and C-peptide were measured using the Elecsys Insulin and C-Peptide assays on a Cobas 801 system (Roche Diagnostics). FFA were measured using the Wako Chemicals NEFA Kit (Fujifilm, Neuss, Germany).

During the clamp trial day, femoral arterial and venous blood samples were drawn into heparinized 2-mL safePICO syringes (Radiometer, Bronshoj, Denmark) for immediate analysis of glucose, lactate, hematocrit, partial pressure of O_2_ (P_O2_) and CO_2_ (P_CO2_), O_2_ saturation, hemoglobin concentration, pH, and bicarbonate ions using an ABL90 Flex blood analyzer (Radiometer). Additional arterial blood was collected as described for the OGTT visit to obtain plasma for subsequent measurements of insulin, FFA, and [6,6-²H₂]glucose enrichment. The enrichment of [6,6-²H₂]glucose in plasma and glucose infusate was quantified using liquid-chromatography tandem mass-spectrometry as previously described^98^.

### Calculations

Muscle glucose uptake and muscle lactate release, as measured by the net exchange of substrates across the leg muscles, were calculated from the femoral arterial plasma flow (F_P_, expressed per kg leg lean mass) and the arteriovenous difference (substrate_a-v_) in plasma concentration of the given substrate, which was corrected for changes in plasma volume by accounting for the net transcapillary water flux (J_v_)^29,99,100^

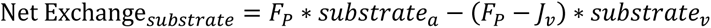

The femoral arterial plasma flow (F_P_) was calculated from the femoral arterial blood flow (F) and the arterial hematocrit (Hct_a_)

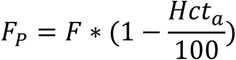

The net transcapillary water flux (J_v_) into or out of the vein was calculated from the femoral arterial blood flow (F), the arterial and venous hemoglobin concentration (Hb_a_ and Hb_v_), and the arterial and venous hematocrit concentration (Hct_a_, and Hct_v_)

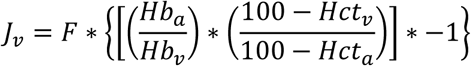

Leg O_2_ consumption (V̇_O2_), leg CO_2_ release (V̇_CO2_) and leg respiratory quotient (RQ) were calculated from the femoral artery blood flow (F) and the arteriovenous difference (Ca-Cv) in content of the given gas

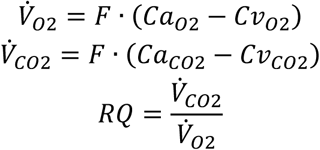

Content of O_2_ and CO_2_ in arterial blood (Ca_O2_ and Ca_CO2_) and venous blood (Cv_O2_ and Cv_CO2_) were computed using the equations by Siggaard-Andersen et al.^101^. Glucose and lipid oxidation across the leg were estimated from the leg V̇_O2_ and V̇_CO2_ using the equations by Péronnet and Massicotte^102^.

Rate of glucose appearance and disappearance were calculated from the [6,6-^2^H_2_]glucose tracer infusion rate and the plasma [6,6-^2^H_2_]glucose enrichment using Steele’s non-steady-state equation^103^. Hepatic glucose production was calculated by subtracting the glucose infusion rate from the [6,6-^2^H_2_]glucose tracer-determined rate of glucose appearance.

The incremental area under the curve (iAUC) for plasma glucose during the OGTT was calculated using the trapezoidal rule. β-cell function was determined by the disposition index, calculated as the product of the early-phase insulinogenic index (IGI) and the insulin sensitivity index (Matsuda index) from the OGTT. Specifically, the early-phase insulinogenic index was calculated as the Δglucose_0-30min_ divided by ΔC-peptide_0-30min_, and the Matsuda index was calculated as 10,000/√(fasting glucose × fasting insulin) × (mean glucose_0–180min_ × mean insulin_0–180min_)^104^.

### Muscle biopsies

Muscle biopsies were obtained from the *vastus lateralis* before initiation (Pre-clamp) and immediately before conclusion (Post-clamp) of the hyperinsulinemic-euglycemic clamp under local anesthesia (10% Lidocaine) using a percutaneous Bergström needle with suction. Upon collection, samples were placed on sterile gauze, rinsed with ice-cold saline to minimize blood contamination, and cleaned of visible fat and connective tissue. The Pre-clamp biopsy was divided into four portions, which were handled differently according to subsequent analyses. One portion of ∼50 mg was snap-frozen in liquid N_2_ for subsequent heteroplasmy, immunoblotting, enzymatic activity, and proteomic analyses. Another portion of ∼15 mg was incubated in 100 mM N-ethylmaleimide (NEM) for 5 min, and snap-frozen in liquid N_2_ for subsequent non-reducing SDS-PAGE and Western blotting. The remaining two portions of ∼90 mg and ∼20 mg were placed in either mitochondrial isolation buffer (100 mM Sucrose, 100 mM KCl, 50 mM Tris-HCl, 1 mM KH2PO4, 0.1 mM EGTA, 0.2% BSA, pH 7.4) or ice-cold biopsy preservation solution (BIOPS: 2.77 mM CaK_2_-EGTA, 7.23 mM K_2_-EGTA, 5.77 mM Na_2_ATP, 6.56 mM MgCl_2_, 20 mM taurine, 15 mM Na_2_Phospho-creatine, 20 mM imidazole, 0.5 mM dithiothreitol, 50 mM K-MES, pH 7.1) and prepared for immediate mitochondrial bioenergetics analyses. The Post-clamp biopsy was snap-frozen in liquid N_2_ for subsequent immunoblotting analyses.

### Muscle mtDNA heteroplasmy

DNA was isolated from ∼5 mg of muscle by QIAamp DNA micro kit (Qiagen, Hilden Germany), according to manufacturer instructions. The entire mtDNA sequence was PCR amplified by AmpliSeq “Precision ID mtDNA Whole Genome Panel” and sequenced on a IonS5 sequencer to a mean depth of >500x by standard protocols (Thermo Fisher Scientific, Waltham, MA). BAM-files were loaded into IGV^105^ and allelic read depth at m.3243 was interpreted as the level of heteroplasmy.

### Muscle mitochondrial bioenergetics

Mitochondrial O_2_ consumption and H_2_O_2_ emission (i.e., production minus removal) rates were measured in permeabilized muscle fibers using high-resolution respirometry and fluorometry (O2k-FluoRespirometer; Oroboros Instruments, Innsbruck, Austria). Additionally, mitochondrial O_2_ consumption rates were measured in isolated mitochondrial fractions using high-resolution respirometry (Oxygraph O2k-FluoRespirometer; Oroboros Instruments).

#### Preparation of permeabilized muscle fibers

Immediately after collection, multiple muscle fiber bundles (1.5-2.5 mg wet weight) were prepared in ice-cold BIOPS by gently teasing the fibers apart with needle tip forceps. Teased fiber bundles were then incubated in BIOPS containing saponin (50 µg/mL) for 30 min under gentle rocking on ice to permeabilize cell membranes while preserving mitochondrial membranes. Thereafter, permeabilized fiber bundles (PmFBs) were rinsed twice for 10 min in 5 mL of mitochondrial respiration medium (MiR05: 0.5 mM EGTA, 3 mM MgCl_2_ · 6H_2_O, 60 mM lactobionic acid, 20 mM taurine, 10 mM KH_2_PO_2_, 20 mM HEPES, 100 mM D-sucrose, and 1 g/L BSA, pH 7.1), gently blotted on dry filter paper, weighed, and transferred to O2k chambers containing 2 mL MiR05.

#### Preparation of isolated mitochondrial fractions

Isolated mitochondrial fractions were prepared according to standard procedure^106^. Briefly, muscle tissue was finely minced for 4 min in ice-cold mitochondrial isolation buffer. After sedimentation and repeated washing steps with mitochondrial isolation buffer, the sample was incubated in mitochondrial isolation buffer containing 0.2 mg/mL bacterial proteinase (Sigma P8038) for 2 min with gentle vortexing. Samples were homogenized using a glass-Teflon pestle for 2 min, diluted in 3 mL of mitochondrial isolation buffer, and centrifuged at 750 *g* for 10 min (4 °C). The supernatant was centrifuged at 10,000 *g* for 10 min, and the resulting pellet washed and re-centrifuged at 7,000 *g* for 4 min. The final mitochondrial pellet was resuspended in suspension buffer (225 mM mannitol, 75 mM Sucrose, 10 mM Tris base, 0.1 mM EDTA, pH 7.4) at 0.6 µL/mg tissue and kept on ice. Aliquots of 5 µL were transferred to O2k chambers containing 2 mL of MiR05. Remaining aliquots were stored at −80 °C.

#### Substrate-inhibitor titration protocol 1

Mitochondrial respiratory capacity in PmFBs was determined using a standardized substrate-inhibitor titration (SUIT) protocol (Fig. 5B-C). First, 5 mM pyruvate, 2 mM malate, and 10 mM glutamate were added to determine leak respiration without adenylates (L_N_). Thereafter, 5 mM ADP was titrated to determine maximal OXPHOS capacity through complex I, followed by titration of 10 mM succinate to determine maximal OXPHOS capacity through complex I and complex II. Then, 10 μM cytochrome C was added to test mitochondrial outer membrane integrity. A stepwise titration of 0.5 μM FCCP (carbonyl cyanide-*p*-trifluoro-methoxyphenyl-hydrazone) was then applied to determine maximal electron transport capacity through complex I and II. Last, 2.5 μM antimycin A was added to inhibit respiration and allow for correction for nonmitochondrial oxygen consumption. For the exercise intervention study, this protocol was further expanded to include titration of 10 mM rotenone to determine electron transport capacity through complex II, followed by titration of 2 mM ascorbate, 0.5 mM TMPD (N,N,N’,N’-Tetramethyl-p-phenylenediamine dihydrochloride), and >100 mM azide to determine electron transport capacity through complex IV. (Fig. 7O). Experiments were performed in MiR05 and at a chamber temperature of 37 °C. O_2_ levels were maintained between 200 and 450 mM to prevent potential O_2_ diffusion limitation.

#### Substrate-inhibitor titration protocol 2

Mitochondrial respiratory capacity and ROS emission capacity in PmFBs were determined using a SUIT protocol validated for simultaneous measurements of mitochondrial O_2_ (*J*O_2_) and H_2_O_2_ (*J*H_2_O_2_) fluxes^29,107^ (Fig. 5G-H and 7R). First, 10 mM succinate was titrated to induce reverse electron flow, followed by titration of 5 mM pyruvate and 0.5 mM malate to stimulate maximal mitochondrial H_2_O_2_ emission. Thereafter, increasing amounts of ADP (25, 100, 175, 5000, and 10000 µM) were titrated stepwise to determine submaximal mitochondrial H_2_O_2_ emission rates. Then, 10 mM glutamate was titrated to determine maximal OXPHOS capacity through complex I and II, followed by titration of 10 µM cytochrome C to test mitochondrial outer membrane integrity. 1 µM oligomycin was then added to inhibit ATP synthase and measure oligomycin-induced leak respiration (LEAK_Omy_). Finally, 2.5 µM antimycin A was titrated to terminate respiration and allow for correction for non-mitochondrial oxygen consumption. Experiments were performed in MiR05 supplemented with 25 µM blebbistatin, 10µM Amplex UltraRed, 5 U/mL superoxide dismutase, and 1 U/mL horseradish peroxidase and at a chamber temperature of 37 °C. Instrumental and chemical O_2_ background fluxes were calibrated as a function of O_2_ concentration and subtracted from the total volume-specific O_2_ flux (Datlab v.7.4 software; Oroboros Instruments). O_2_ levels were maintained between 200 and 450 mM. Standardized instrumental and chemical calibrations were conducted according to the manufacturer’s instructions (Oroboros Instruments, Innsbruck, Austria). These allowed for corrections for i) background diffusion of O_2_ into the chamber, ii) O_2_ solubility in MiR05, and iii) background consumption of O_2_ by the electrodes, which was determined across the range of chamber O_2_ concentrations employed during the experiments (200–450 mM).

#### Substrate-inhibitor titration protocol 3 and 4

Mitochondrial respiratory capacity in isolated mitochondria was determined using a carbohydrate- and a fatty acid-supported SUIT protocol (Fig. 5L, N, P). The carbohydrate-supported protocol started with titration of 5 mM pyruvate and 10 mM malate, followed by 5 mM ADP, 10 mM glutamate, 10 mM succinate, and 1 μM oligomycin. The fatty acid-supported protocol started with titration of 10 μM palmitoyl carnitine and 2 mM malate, followed by 5 mM ADP, 5 mM pyruvate, 10 mM glutamate, 10 mM succinate, and 1 μM oligomycin. Experiments were performed in MiR05 and at a chamber temperature of 37 °C. O_2_ levels were maintained between 0 and 200 mM.

#### Compounds and substrates

The following compounds and substrates were used to evaluate mitochondrial bioenergetics: Amplex UltraRed (Life Technologies, A36006); superoxide dismutase (Sigma-Aldrich, S 8160); blebbistatin (Sigma-Aldrich, B 0560); horseradish peroxidase (Sigma-Aldrich, P 8250); pyruvate (Sigma-Aldrich, P 2256); malate (Sigma-Aldrich, M 1000); glutamate (Sigma-Aldrich, G 1626); ADP (Sigma-Aldrich, A 5285); succinate (Sigma-Aldrich, S 2378); FCCP (Sigma-Aldrich, C 2920); rotenone (Sigma-Aldrich, R8875); ascorbate (Sigma-Aldrich, A 7631); TMPD (Sigma-Aldrich, T 3134); azide (Sigma-Aldrich, S 2002); palmitoyl carnitine (Sigma-Aldrich, P 4509); cytochrome C (Sigma-Aldrich, C 7752); oligomycin (Sigma-Aldrich, O 4876); and antimycin A (Sigma-Aldrich, A 8674).

### Mitochondrial enzymatic activity

Citrate synthase (CS) maximal activity was quantified in both muscle homogenates and isolated mitochondrial fractions as described previously^108^. CS activity was used as a marker of mitochondrial content^44^ and for normalization of mitochondrial O_2_ fluxes.

### Immunoblotting

Protein abundance in whole-muscle homogenate lysates was determined by SDS-PAGE and Western blotting. Protein markers of muscle subcellular redox state (peroxiredoxin dimerization) were resolved by SDS-PAGE under non-reducing conditions^109^.

#### Whole-muscle lysate preparation, SDS-PAGE and Western Blotting

Freeze-dried muscle samples were dissected under a stereomicroscope to remove visible blood, fat, and connective tissue. Dissected muscle tissue was weighed, and a fresh batch of ice-cold homogenization buffer (10% glycerol, 20 mM Na-pyrophosphate, 150 mM NaCl, 50 mM HEPES (pH 7.5), 1% Nonidet P-40 (NP-40), 20 mM β-glycerophosphate, 2 mM Na3VO4, 10 mM NaF, 2 mM PMSF, 1 mM EDTA (pH 8), 1 mM EGTA (pH 8), 10 µg/mL aprotinin, 10 µg/mL leupeptin, and 3 mM benzamidine) was added. For samples designated for non-reducing SDS-PAGE, 100 mM NEM was included in the homogenization buffer. Approximately 2 mg (dry weight) of muscle tissue was homogenized for 2 min at 30 Hz in a TissueLyser II (Qiagen, Retsch GmbH, Haan, Germany). Homogenates were then rotated for 1 h at 4 °C, followed by centrifugation at 17,500 g for 20 min at 4°C. The supernatant (lysate) was collected, and the total protein concentration in each lysate sample was determined in triplicate using a BSA-based assay (Thermo Scientific). Each lysate sample was then mixed with 6×Laemmli buffer (7 mL of 0.5 M Tris base, 3 mL of glycerol, 0.93 g of DTT, 1 g of SDS, and 1.2 mg of bromophenol blue) and double-distilled H_2_O to achieve a final protein concentration of 2.0 µg/µL. For non-reducing SDS-PAGE, DTT was omitted from the Laemmli buffer.

Equal amounts of total protein were loaded onto precast Criterion TGX Stain-Free gels (Bio-Rad, Copenhagen, Denmark), with all samples from i) a given pair of matched carriers and controls or ii) the non-trained and trained leg from the same participant run on the same gel. An internal standard was included on each gel to allow comparison across blots. Proteins were separated by SDS-PAGE and semi-dry transferred to a PVDF membrane (Millipore, Denmark). Membranes were blocked in either 2% skim milk or 3% BSA prepared in

Tris-buffered saline-Tween 20 (TBST), followed by overnight incubation at 4 °C with primary antibody diluted in the corresponding blocking solution. After TBST washes, membranes were incubated for 1 h at room temperature with a horseradish peroxidase-conjugated secondary antibody (1:5,000 dilution in 2% skim milk or 3% BSA). Membrane staining was visualized by incubation with a chemiluminescent horseradish peroxidase substrate (Immobilon Forte, Millipore, Denmark) before image digitalization on ChemiDoc MP (Bio-Rad). Western blot band intensities were quantified by densitometry (total band intensity adjusted for background intensity) using GelGenie^110^. For signaling proteins, phosphorylated and total protein abundances were quantified on separate membranes in independent analyses, and none of the membranes were stripped prior to quantification.

#### Antibodies

The following antibodies were used: Phospho-Akt (Thr308) (Cell Signaling, catalog no. 9275); Phospho-Akt (Ser473) (Cell Signaling, catalog no. 9271); Akt2 (Cell Signaling, catalog no. 3063); Phospho-GSK-3β (Ser9) (Cell Signaling, catalog no. 5558); GSK-3β (BD Transduction Laboratories, catalog no. 610202); Phospho-TBC1D4 (Thr642) (Cell Signaling, catalog no. 8881); TBC1D4 (Abcam, catalog no. ab189890); Phospho-p70S6K (Thr389) (Cell Signaling, catalog no. 9205); p70S6K (Cell Signaling, catalog no. 9202); Phospho-AMPKα2 (Thr172) (Cell Signaling, catalog no. 2531); AMPKα2 (Abcam, catalog no. 3750); PRDX2 (Abcam, catalog no. ab109367); PRDX3 (Abcam, catalog no. ab73349).

### Muscle proteomics

#### Sample preparation

Freeze-dried skeletal muscle samples were dissected free of visible blood, fat, and connective tissue. Approximately 4 mg of dry tissue was transferred to Micronic tubes containing a metallic bead, submerged in liquid nitrogen, and homogenized using a 1600 MiniG SPEX SamplePrep instrument (3 × 30 s at 1,500 rpm, with re-freezing in liquid N_2_ between cycles). Homogenized tissue was lysed in 200 µL lysis buffer (1% [w/v] SDC, 100 mM Tris-HCl, pH 8.5) supplemented with nuclease (1 µL per 50 µL buffer). Samples were boiled at 95 °C for 10 min at 1,000 rpm (Thermomixer), sonicated (15 min, 30 s on/off, maximum power), and further homogenized using a BeatBox (Preomics, Germany) at high setting for 3 × 10 min. Lysates were centrifuged (12,000 *g* for 10 min) and the supernatant collected.

Protein concentration was determined by BCA assay using a standard curve, and 100 µg total protein was used for reduction, alkylation, and enzymatic digestion. Extracted proteins were reduced with 5 mM tris(2-carboxyethyl)phosphine (TCEP) for 15 min at 55 °C, alkylated with 20 mM chloroacetamide (CAA) for 30 min at room temperature, and digested with Trypsin/LysC (1:50 enzyme-to-protein ratio). Peptides were cleaned up using iST-PSI desalting plates (PreOmics) and dried by vacuum centrifugation. Dried peptides were resuspended in buffer A* (0.1% trifluoroacetic acid, 2% acetonitrile) prior to LC-MS analysis.

#### Liquid chromatography mass spectrometry

Peptide separation was performed on an Aurora (Gen3) 25 cm, 75 μM ID column packed with C18 beads (1.6 μm) (IonOpticks) using a Vanquish Neo UHPLC system (Thermo Fisher Scientific). A 60-min stepped gradient was employed as follows: 2-17% solvent B (0.1% formic acid in acetonitrile) for 33 min, 17-25% solvent B for 11 min, and 25-35% solvent B for 6 min, at a constant flow rate of 400 nL/min and column temperature of 50 °C. Eluted peptides were injected via a Nanospray Flex ion source into a Tribrid Ascend mass spectrometer (Thermo Fisher Scientific). Spray Voltage was set to 2200 (V). Data was acquired in data-independent acquisition (DIA) mode with the Orbitrap MS resolution 60,000 (400-900 m/z range), AGC target 250%, and maximum injection time 123 ms. DIA scans (41 scans, 12 Th width, 1 Th overlap) were acquired at 15,000 resolution, AGC target 1000%, and maximum injection time 27 ms, using HCD fragmentation normalized collision energy (NCE) of 30%.

#### Data processing and bioinformatics analysis

Raw MS files were processed using DIA-NN (v.2.2.0) in directDIA mode with a spectral library predicted from the human UniProt FASTA database (81,387 protein isoforms). Highly heuristic protein grouping and Match Between Runs were enabled. Carbamidomethylation of cysteine was set as a fixed modification, and oxidation of methionine, acetylation at the protein N-terminus, and N-terminal methionine excision as variable modifications. Up to one missed cleavage and a maximum of two variable modifications per peptide were allowed. The minimum peptide amino acid length was 7. The protein groups and precursors were filtered at 1% FDR. All other settings were set as default.

Quantified protein intensities were log_2_-transformed, and features with less than at least 70% valid values in at least one group were filtered out. Missing value were imputed using Mixed imputation, consisting of imputing values missing at random with kNN, and values missing not at random with MinProb^111^. Quantified protein intensities were median-normalized and statistical analyses and data visualization were performed in R v.4.3.1 (https://www.r-project.org/). Principal component analysis was conducted on the complete quantified protein matrix.

#### Differential abundance analysis

Differential protein abundance was assessed using the *limma* package with linear modeling (*lmFit*) and eBayes smoothening^112^. For the cross-sectional study, the model included group (carriers vs. controls) as a factor, age as a covariate, and matched pairs as a blocking variable. For the exercise intervention study, the model included leg (trained vs. non-trained) as a factor, muscle heteroplasmy as a covariate, and participants as a blocking variable. Proteins with a Benjamini-Hochberg (BH) corrected *P* value < 0.05 (5% FDR) were considered significantly differentially abundant.

#### Global muscle proteome pathway enrichment analyses

Gene Ontology (GO)-based pathway enrichment analysis was performed using the *clusterProfiler* package^113^. Redundant GO Biological Process and Cellular Component terms with semantic similarity > 70% were filtered to retain only the most significant representative term. BH-corrected *P* values < 0.05 were considered significant.

#### Within-pair correlation-based pathway enrichment analyses

Within-pair correlation-based pathway enrichment analysis was conducted using proteomics and muscle insulin sensitivity data from individually matched carriers and controls. For each protein, within-matched carrier-control pair differences in abundance (carrier – control) were correlated with within-pair differences in insulin-stimulated leg glucose uptake using Kendall’s rank correlation (τ). GO-based enrichment analysis was then performed as described above.

#### Oxidative, glycolytic, and mitochondrial protein enrichment

Oxidative metabolism proteins were defined as those annotated in *GO Biological Process*, *Reactome*, or *KEGG* with the terms “tricarboxylic acid cycle”, or “TCA cycle”, or “TCA”, or “electron transport chain”, or “oxidative phosphorylation”. Glycolytic metabolism proteins were defined as those annotated with the terms “glycolysis” or “glycolytic process” in any of the same resources. Mitochondrial proteins were identified using MitoCarta3.0^40^. For each sample, log_2_ intensities were back-transformed to linear scale (raw intensity), and the relative abundance of oxidative, glycolytic, and mitochondrial proteins was computed as the summed raw intensity of oxidative, glycolytic, and mitochondrial proteins relative to the summed raw intensity of the total proteome.

#### Mitochondrial proteome-specific pathway enrichment analysis

Group-based pathway enrichment analysis was performed on MitoCarta3.0 “MitoPathways” using the *clusterProfiler* package^113^. BH-corrected P values < 0.05 were considered significant.

To assess MitoPathways enrichment at the individual level, single-sample (ss) enrichment analysis was applied using the *GSVA* package^114^. Per-sample enrichment scores for MitoPathways were summarized within matched carrier-control pairs as Δ (m.3243A>G – Controls) enrichment scores for the cross-sectional study, and within-participant as Δ (trained leg – non-trained leg) enrichment scores for the exercise intervention study.

#### Functional classification of mitochondrial bioenergetic and non-bioenergetic proteins

Mitochondrial proteins were manually grouped into three broad functional classes based on their MitoCarta3.0 MitoPathways (Tier 3) annotations. “Core bioenergetic” proteins were defined as those participating directly in oxidative metabolism and ATP-producing pathways, including TCA cycle, fatty acid and amino acid oxidation, pyruvate and ketone metabolism, the malate-aspartate and glycerol phosphate shuttles, and electron transport chain complexes I-V and their assembly factors. “Enabling bioenergetic” proteins comprised pathways that support or regulate mitochondrial energy metabolism, such as cofactor and coenzyme biosynthesis (heme, coenzyme Q, NAD⁺, Fe-S clusters), mitochondrial carriers and transporters, phospholipid and cardiolipin metabolism, and calcium handling. “Non-bioenergetic” proteins included all remaining MitoPathways not directly involved in energy metabolism, such as those related to mitochondrial protein import, translation, dynamics, and quality control.

#### Normalization to mitochondrial enrichment

To account for bias arising from between-group and between-leg differences in total mitochondrial protein abundance and to enable detection of non-stoichiometric alterations within the mitochondrial proteome (i.e., relative differences in the mitochondrial proteome composition independent of total mitochondrial content), protein intensities were normalized to mitochondrial protein enrichment following the BESt-normalization approach^41,42^. Specifically, variance-stabilizing normalization (VSN) was applied across the mitochondrial protein subset, after which differential abundance analysis and MitoPathways enrichment analysis were performed on the normalized mitochondrial proteome as described for the global muscle proteome.

### Statistical analysis

#### Cross-sectional study

Sample size calculations were based on a prior cross-sectional study of m.3243A>G carriers and healthy controls^17^, which reported large effect sizes for whole-body insulin sensitivity (*d* = 1.8) and muscle insulin sensitivity (*d* = 1.7) under a two-sided α = 0.05, corresponding to power >0.98 with group sizes of n=14 and n=13, respectively. Anticipating smaller effects for other primary outcomes (muscle mitochondrial function and proteomic signatures), we implemented an individually matched design (age, sex, BMI, and physical activity) to increase power by controlling for major confounders. Based on this design, the target sample size was 15 carriers and 15 controls.

Participants were recruited in matched pairs (carrier-control). Primary analyses used linear mixed-effects models (LMMs) with group (carrier vs. control) as a fixed effect and a random intercept for matched pair. Specifically, for comparisons of participant characteristics (Table S1), LMMs were: *outcome ∼ group + (1 | matched pair)*; for outcomes related to the HE clamp (Fig. 1 and S2), OGTT (Fig. S3), proteome signatures (Fig. 3 and 4), and mitochondrial bioenergetics (Fig. 5), LMMs were: *outcome ∼ group + age + (1 | matched pair)*; for outcomes related to insulin signaling (Fig. 2), LMMs were: *phosphoprotein_abundance ∼ group * condition + age + total_protein_abundance + (1 | matched pair)*.

Secondary analyses used linear regression models adjusted for age, sex, BMI, and physical activity to explore associations between muscle mtDNA heteroplasmy and outcome measures related to glucose homeostasis and mitochondrial characteristics (Fig. S4).

#### Exercise intervention study

Sample size calculations were based on a prior study with a similar within-participant design^115^, which reported a large effect size for muscle insulin sensitivity in the trained versus non-trained leg (*d* = 1.4) under a two-sided α = 0.05, corresponding to power >0.97 with a sample size of n=10. LMMs included leg (trained vs. non-trained) as a fixed effect, heteroplasmy as a covariate, and a random intercept for participant. Specifically, for outcomes related to the HE clamp (Fig. 6D, F, H), proteome signatures (Fig. 7F, G, J, K, and L), and mitochondrial bioenergetics (Fig. 7P, Q, and S), LMMs were: *outcome ∼ leg + heteroplasmy + (1 | participant)*; for outcomes related to insulin signaling (Fig. 6J-N), LMMs were: *phosphoprotein_abundance ∼ leg * condition + heteroplasmy + total_protein_abundance + (1 | participant)*. *Cross-study integration and association analyses*

To test whether exercise reverses proteomic defects identified cross-sectionally, study-specific *limma* results were converted to standardized z-scores that encode both the direction and statistical confidence of each protein’s differential abundance, thereby enabling direct comparison across studies (Fig. 8A). These z-scores were combined across studies using Stouffer’s method^116^ to yield a per-protein rescue statistic that is large when the exercise effect directionally opposes the m.3243A>G variant effect. Two-sided BH-corrected *P* values were derived. Proteins with opposite-sign effects across studies and rescue FDR<0.05 were classified as exhibiting consistent rescue. For pathway-level analyses (GO-BP, GO-CC, and MitoCarta-MitoPathways), per-sample pathway scores were computed as the mean of row-z-scored protein abundances across pathway members within each study’s median-normalized expression matrix, and the resulting score matrices were analyzed using the same *limma* models described above. Cross-study rescue statistics and Spearman correlations between negated cross-sectional and exercise effects were computed at each level.

Exploratory analyses were performed to examine associations of pre-specified molecular signatures with insulin-stimulated leg glucose uptake across the cross-sectional and exercise cohorts (Fig. 8D). Given the paired structure of both studies, within-cluster associations were estimated using study-specific fixed-effects models: each molecular signature was standardized within cohort (z-scored) and the model was specified as: *leg glucose uptake ∼ molecular signature + cluster*, where cluster denotes matched pairs (m.3243A>G vs. Controls) in the cross-sectional cohort and participants (trained vs. non-trained leg) in the exercise cohort. To obtain an overall estimate across studies (pooled β), the two study-specific within-cluster β coefficients and their standard errors were combined using inverse-variance fixed-effect meta-analysis (*metafor* package). The same framework was applied to assess associations between PRKAG2 abundance and MitoPathways scores, with FDR correction across all tested pathways.

#### Common procedures

The level of significance for all analyses was set at *P*<0.05. For LMM-based analyses, *P*-values were computed using Kenward-Roger approximation of the degrees of freedom. Model diagnostics were based on visual inspection of residual plots, supported by Shapiro-Wilk tests. In case of heteroscedasticity, outcomes were log-transformed prior to analysis and back-transformed for presentation as geometric means with 95% confidence intervals (CIs). Analyses were performed with R v.4.3.1 (https://www.r-project.org/). Key packages included *lme4* and *emmeans* for linear mixed-effects modelling and *limma* for differential protein abundance. Data are graphically presented as observed individual values overlaid with model-estimated means and 95% CIs, unless otherwise stated.

## SUPPLEMENTARY MATERIAL

**Table S1.** Participant characteristics.

|  | Cross-sectional cohort |  | Exercise cohort |
| --- | --- | --- | --- |
|  | m.3243A>G | Controls | m.3243A>G |
| Sex (female/male) | 11/4 | 11/4 | 5/6 |
| Age (years) | 45.3 ± 12.9 | 45.5 ± 12.8 | 45.5 ± 11.7 |
| Sedentary time (min/day) | 696 ± 74 | 690 ± 92 | N/A |
| LPA time (min/day) | 165 ± 40 | 164 ± 33 | N/A |
| MVPA time (min/day) | 50 ± 39 | 53 ± 37 | N/A |
| Height (cm) | 169 ± 6 | 171 ± 8 | 171 ± 9 |
| Weight (kg) | 67.4 ± 13.3 | 73.9 ± 15.5 | 70.3 ± 19.3 |
| Body mass index (kg/m<sup>2</sup>) | 23.6 ± 4.2 | 25.0 ± 4.4 | 23.8 ± 4.9 |
| Waist-to-hip ratio | 0.83 ± 0.13 | 0.81 ± 0.10 | 0.81 ± 0.14 |
| Fat mass (kg) | 21.8 ± 8.4 | 23.5 ± 7.7 | 21.0 ± 12.1 |
| Fat-free mass (kg) | 42.8 ± 9.1* | 47.5 ± 10.7 | 46.6 ± 12.2 |
| Percent body fat mass (%) | 33 ± 10 | 33 ± 8 | 30 ± 12 |
| Percent body fat-free mass (%) | 64 ± 9 | 64 ± 7 | 67 ± 12 |
| **Fasting plasma parameters** |  |  |  |
| Glucose (mM) | 5.9 ± 1.6* | 4.9 ± 0.4 | 6.1 ± 1.2 |
| Insulin (pM) | 76 ± 51* | 42 ± 14 | 49.7 ± 37 |
| Insulin C-peptide (pM) | 780 ± 366* | 510 ± 110 | 624 ± 352 |
| Hemoglobin A1c (mmol/mol) | 37.9 ± 6.0* | 33.9 ± 3.2 | 39.0 ± 6.8 |
| HOMA-β (%) | 112 ± 52 | 108 ± 20 | 92 ± 30 |
| HOMA-S (%) | 106 ± 73 | 139 ± 59 | 110 ± 74 |
| HOMA2-IR | 1.70 ± 1.19* | 0.90 ± 0.30 | 1.34 ± 0.83 |
| Total cholesterol (mM) | 5.0 ± 1.3 | 4.7 ± 0.6 | 4.8 ± 1.1 |
| HDL cholesterol (mM) | 1.6 ± 0.5 | 1.7 ± 0.4 | 1.5 ± 0.4 |
| LDL cholesterol (mM) | 3.1 ± 1.0 | 2.9 ± 0.6 | 3.1 ± 0.9 |
| Triglycerides (mM) | 1.3 ± 1.0* | 0.8 ± 0.4 | 1.0 ± 0.5 |
| Alanine aminotransferase (IU/L) | 22.8 ± 10.2 | 20.5 ± 20.4 | 18.7 ± 13.0 |
| Lactate (mM) | 1.16 ± 0.54* | 0.78 ± 0.32 | 1.17 ± 0.60 |
| Creatine kinase (U/L) | 73 ± 23 | 96 ± 52 | 88 ± 37 |
| **Cardiovascular and respiratory parameters** |  |  |  |
| Systolic blood pressure (mmHg) | 118 ± 13* | 110 ± 11 | 117.4 ± 13.0 |
| Diastolic blood pressure (mmHg) | 76 ± 9* | 70 ± 9 | 74.0 ± 13.0 |
| Resting heart rate (bpm) | 66 ± 15 | 60 ± 9 | 70 ± 13 |
| VO<sub>2</sub>peak (mL/min/kg) | 29.2 ± 6.4* | 36.7 ± 8.6 | 30.1 ± 5.9 |
| **Medications** |  |  |  |
| Thyroid hormone supplements | - | 1 | - |
| Hormone IUD | 1 | 3 | - |
| Cholesterol absorption inhibitors | 1 | - | - |
| Angiotensin II -receptor blocker | 1 | - | - |
| ACE-inhibitors | 1 | - | - |
| Calcium channel blocker | 1 | - | - |
| 5-ASA | 1 | - | 1 |
| Sulfonylurea | 1 | - | 1 |
| Insulin | 2 | - | 4 |
| Oral contraceptives | 2 | - | 1 |

LPA, light intensity physical activity; MVPA, moderate-to-vigorous intensity physical activity; HOMA-β, homeostatic model assessment of β-cell function; HOMA-S, homeostatic model assessment of insulin sensitivity; HOMA2-IR, homeostatic model assessment of insulin resistance <sup>2118</sup>; 5-ASA, 5-amino salicylic acid; HDL/LDL, high-/low-density lipoprotein; VO<sub>2</sub>peak, peak oxygen uptake; ACE, angiotensin-converting enzyme; IUD, intrauterine device. Data are presented as means ± SD. \*Different from Controls (P< 0.05).

**Table S2.**
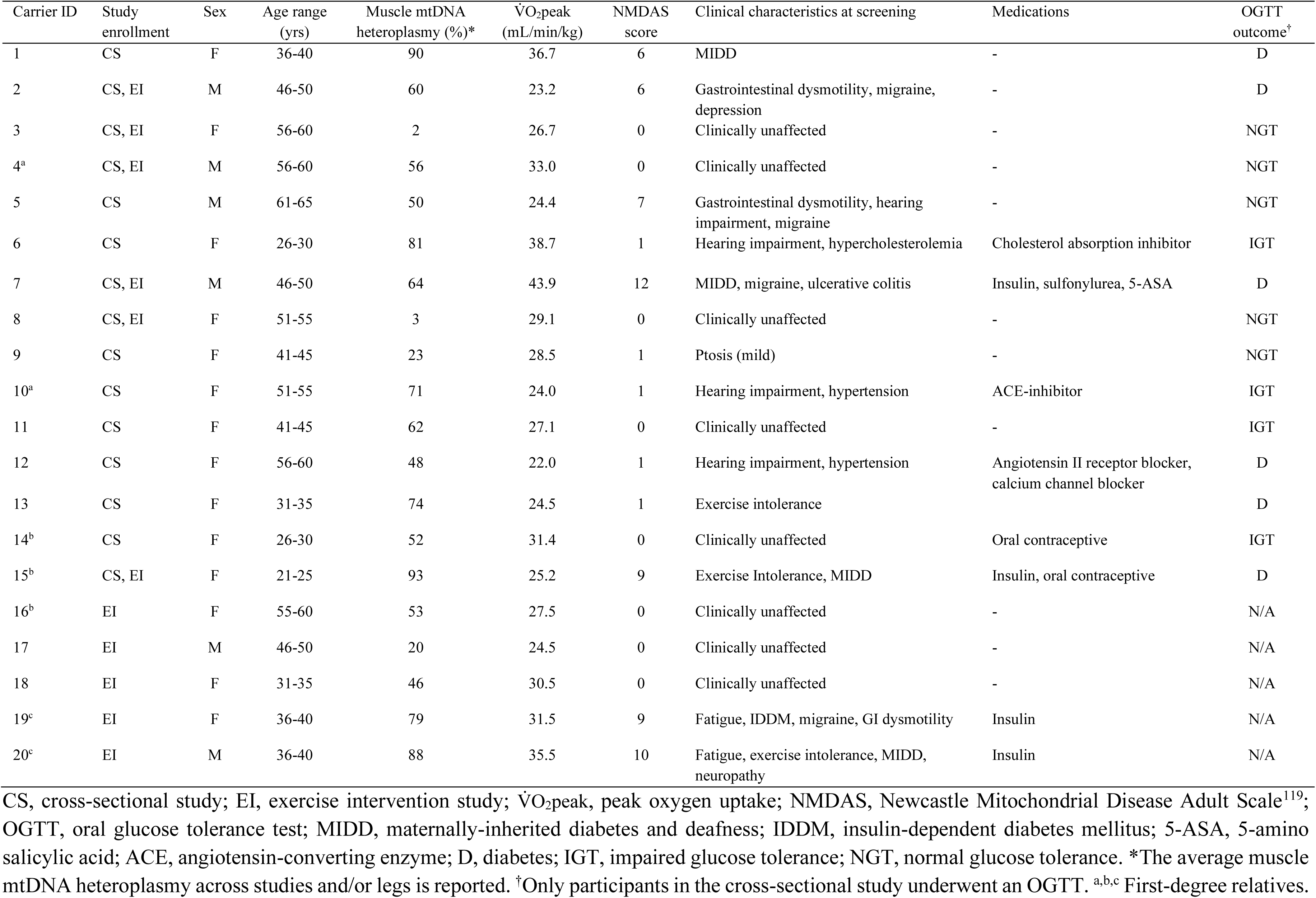
Demographic and clinical characteristics of m.3243A>G carriers.

| Carrier ID | Study enrollment | Sex | Age range (yrs) | Muscle mtDNA heteroplasmy (%)* | $\dot{V}O_2$peak (mL/min/kg) | NMDAS score | Clinical characteristics at screening | Medications | OGTT outcome<sup>†</sup> |
| --- | --- | --- | --- | --- | --- | --- | --- | --- | --- |
| 1 | CS | F | 36-40 | 90 | 36.7 | 6 | MIDD | - | D |
| 2 | CS, EI | M | 46-50 | 60 | 23.2 | 6 | Gastrointestinal dysmotility, migraine, depression | - | D |
| 3 | CS, EI | F | 56-60 | 2 | 26.7 | 0 | Clinically unaffected | - | NGT |
| 4<sup>a</sup> | CS, EI | M | 56-60 | 56 | 33.0 | 0 | Clinically unaffected | - | NGT |
| 5 | CS | M | 61-65 | 50 | 24.4 | 7 | Gastrointestinal dysmotility, hearing impairment, migraine | - | NGT |
| 6 | CS | F | 26-30 | 81 | 38.7 | 1 | Hearing impairment, hypercholesterolemia | Cholesterol absorption inhibitor | IGT |
| 7 | CS, EI | M | 46-50 | 64 | 43.9 | 12 | MIDD, migraine, ulcerative colitis | Insulin, sulfonylurea, 5-ASA | D |
| 8 | CS, EI | F | 51-55 | 3 | 29.1 | 0 | Clinically unaffected | - | NGT |
| 9 | CS | F | 41-45 | 23 | 28.5 | 1 | Ptosis (mild) | - | NGT |
| 10<sup>a</sup> | CS | F | 51-55 | 71 | 24.0 | 1 | Hearing impairment, hypertension | ACE-inhibitor | IGT |
| 11 | CS | F | 41-45 | 62 | 27.1 | 0 | Clinically unaffected | - | IGT |
| 12 | CS | F | 56-60 | 48 | 22.0 | 1 | Hearing impairment, hypertension | Angiotensin II receptor blocker, calcium channel blocker | D |
| 13 | CS | F | 31-35 | 74 | 24.5 | 1 | Exercise intolerance | - | D |
| 14<sup>b</sup> | CS | F | 26-30 | 52 | 31.4 | 0 | Clinically unaffected | Oral contraceptive | IGT |
| 15<sup>b</sup> | CS, EI | F | 21-25 | 93 | 25.2 | 9 | Exercise Intolerance, MIDD | Insulin, oral contraceptive | D |
| 16<sup>b</sup> | EI | F | 55-60 | 53 | 27.5 | 0 | Clinically unaffected | - | N/A |
| 17 | EI | M | 46-50 | 20 | 24.5 | 0 | Clinically unaffected | - | N/A |
| 18 | EI | F | 31-35 | 46 | 30.5 | 0 | Clinically unaffected | - | N/A |
| 19<sup>c</sup> | EI | F | 36-40 | 79 | 31.5 | 9 | Fatigue, IDDM, migraine, GI dysmotility | Insulin | N/A |
| 20<sup>c</sup> | EI | M | 36-40 | 88 | 35.5 | 10 | Fatigue, exercise intolerance, MIDD, neuropathy | Insulin | N/A |

CS, cross-sectional study; EI, exercise intervention study; $\dot{V}O_2$ peak, peak oxygen uptake; NMDAS, Newcastle Mitochondrial Disease Adult Scale<sup>119</sup>; OGTT, oral glucose tolerance test; MIDD, maternally-inherited diabetes and deafness; IDDM, insulin-dependent diabetes mellitus; 5-ASA, 5-amino salicylic acid; ACE, angiotensin-converting enzyme; D, diabetes; IGT, impaired glucose tolerance; NGT, normal glucose tolerance. \*The average muscle mtDNA heteroplasmy across studies and/or legs is reported. <sup>†</sup>Only participants in the cross-sectional study underwent an OGTT. <sup>a,b,c</sup> First-degree relatives.

**Figure S1.**
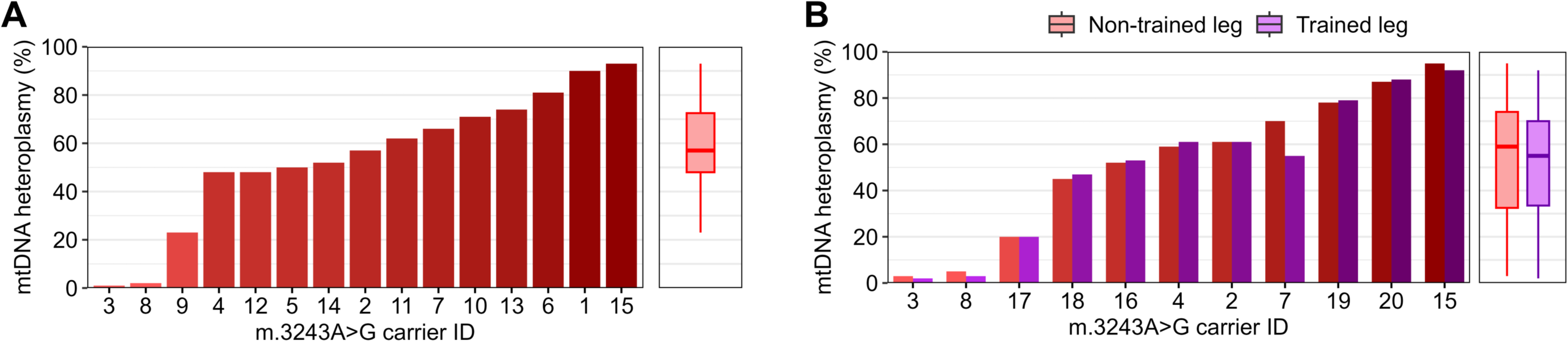
Skeletal muscle mtDNA heteroplasmy in m.3243A>G carriers. (A) Muscle mtDNA heteroplasmy in m.3243A>G carriers enrolled in the cross-sectional study. (B) Muscle mtDNA heteroplasmy in non-trained and trained leg from m.3243A>G carriers enrolled in the exercise intervention study.

**Figure S2.**
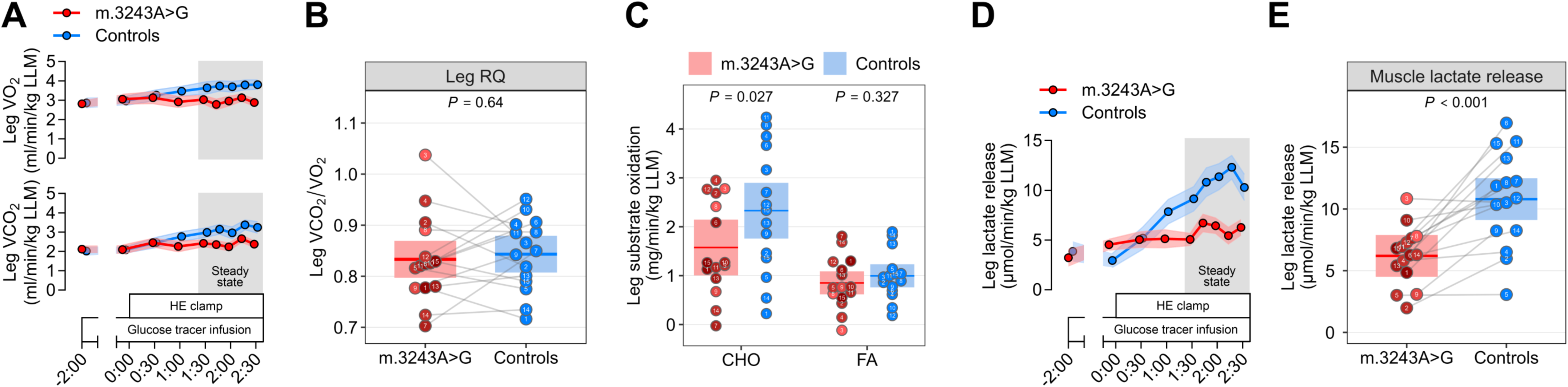
Reduced muscle carbohydrate oxidation and lactate release in m.3243A>G carriers under hyperinsulinemic-euglycemic conditions. (A) Time course of leg O_2_ consumption (V̇O_2_) and CO_2_ release (V̇CO_2_) (data are means ± SEM). (B) Respiratory quotient (RQ) during the steady-state period of the hyperinsulinemic-euglycemic (HE) clamp. (C) Carbohydrate (CHO) and fatty acid (FA) oxidation across the leg, as estimated from the leg V̇O_2_ and V̇CO_2_, during the steady-state period of the HE clamp. (D and E) Leg lactate release as calculated from the arteriovenous difference in plasma lactate concentration and the femoral arterial blood flow. [(D)] Time course of leg lactate release (data are means ± SEM). [(E)]) Leg lactate release during the steady-state period of the HE clamp. Linear mixed models were used to estimate between-group differences [(B), (C), and (E)]. Data are presented as observed individual values (with lines connecting individually matched participants) and estimated means ± 95% confidence limits, unless otherwise stated. For m.3243A>G carriers, individual datapoints are color-shaded to indicate muscle mtDNA heteroplasmy (light red = low, dark red = high). n = 30.

**Figure S3.**
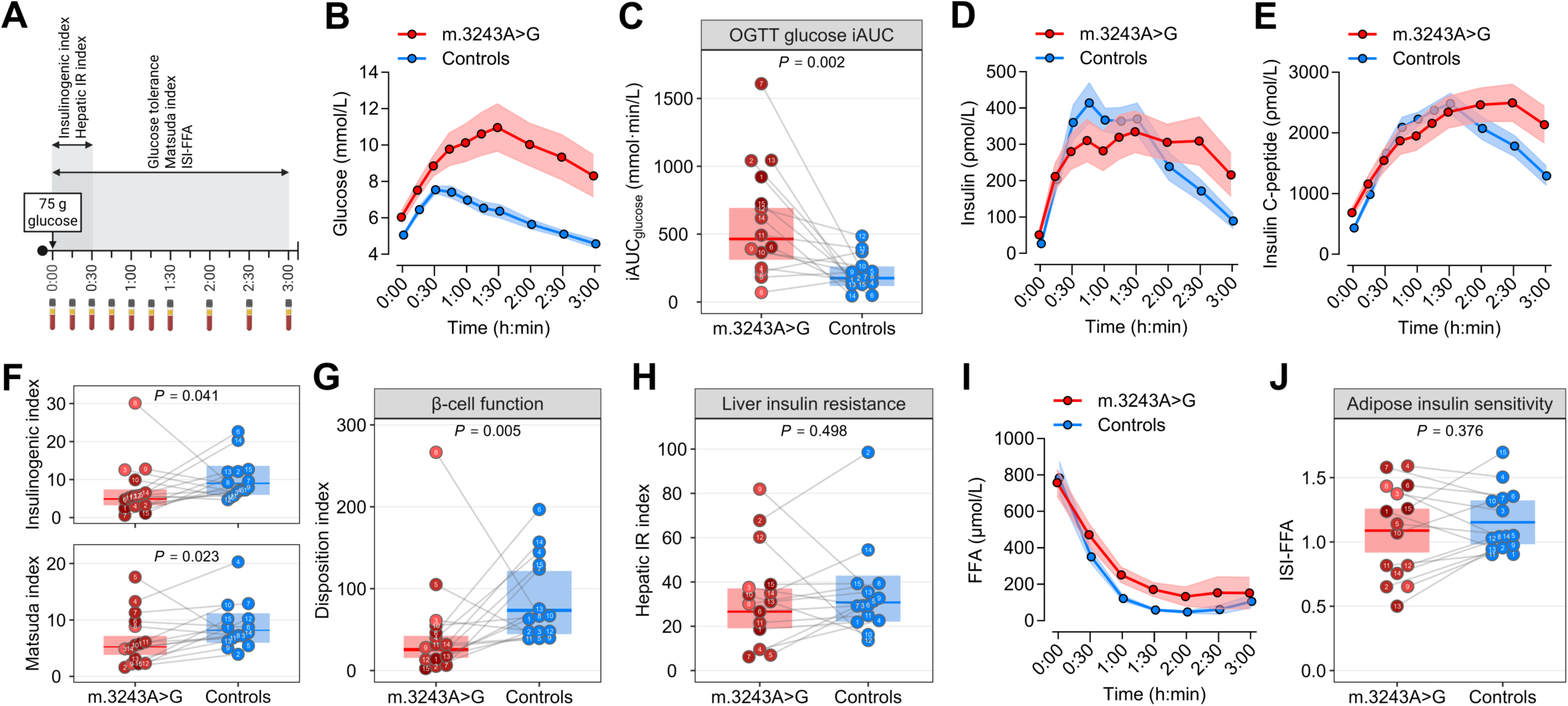
Impaired glucose tolerance due to concomitant defects in β-cell function and systemic insulin sensitivity in m.3243A>G carriers. (A) Workflow of the oral glucose tolerance test (OGTT) visit, including timeframes for each outcome measure. (B and C) Glucose tolerance as measured by the incremental area under the curve (iAUC) of plasma glucose during the OGTT. [(B)] Time course of plasma glucose (data are means ± SEM). (D and E) Time course of plasma insulin and insulin C-peptide during the OGTT (data are means ± SEM). (F) Early-phase insulinogenic index (IGI) and insulin sensitivity index (Matsuda index) from the OGTT. (G) β-cell function expressed as the disposition index, calculated as the product of IGI and the Matsuda index from the OGTT. (H) Liver insulin resistance, calculated as the hepatic IR index based on plasma glucose and insulin during the first 30 min of the OGTT^120^. (I-J) Adipose tissue insulin sensitivity, calculated as the insulin sensitivity index for plasma FFA (ISI-FFA)^121^. [(I)] Time course of plasma FFA (data are means ± SEM) Linear mixed models were used to estimate between-group differences [(E), (F), (G), and (H)]. Data are presented as observed individual values (with lines connecting individually matched participants) and estimated means ± 95% confidence limits, unless otherwise stated. For m.3243A>G carriers, individual datapoints are color-shaded to indicate muscle mtDNA heteroplasmy (light red = low, dark red = high). n = 30.

**Figure S4.**
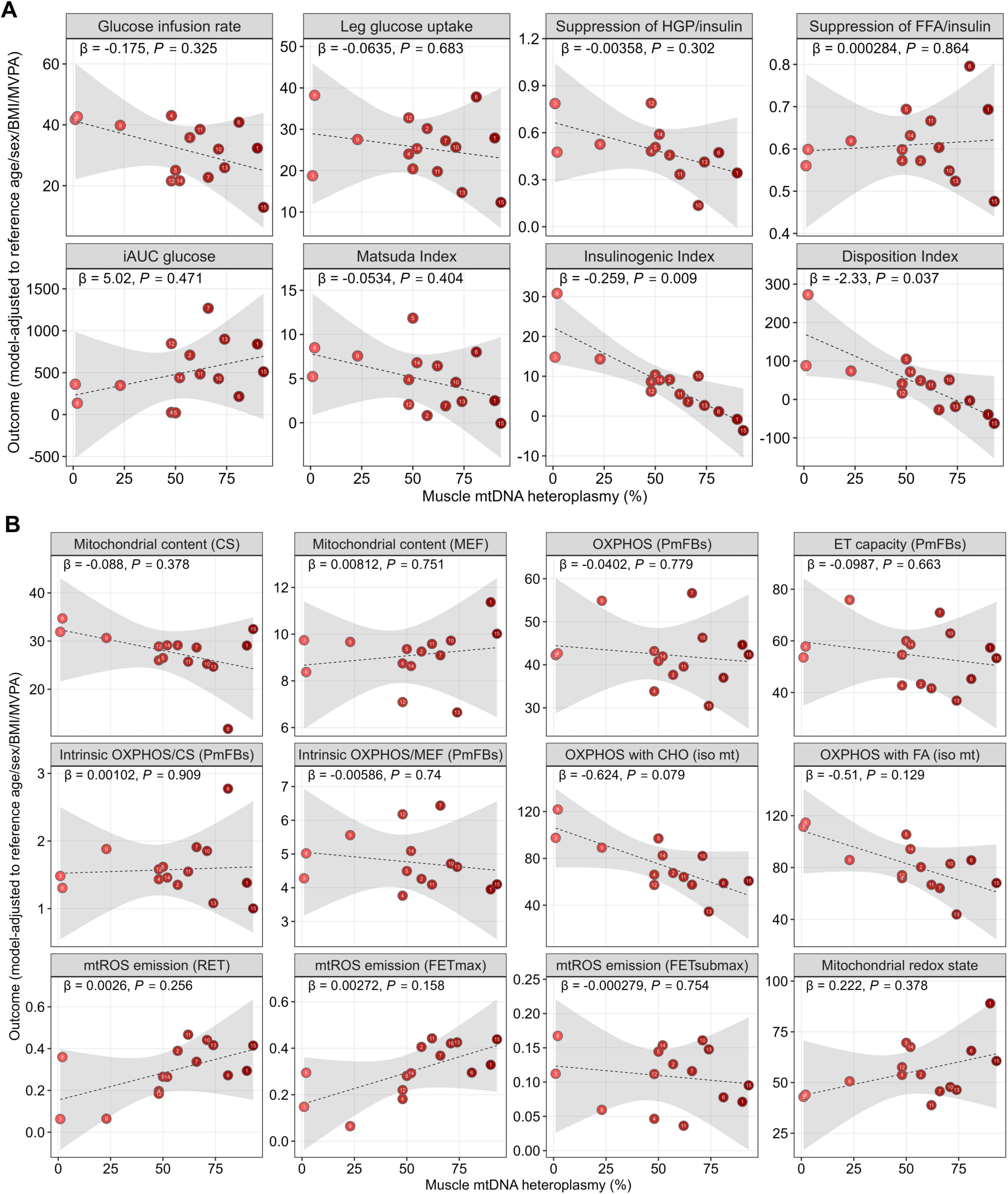
Skeletal muscle mtDNA heteroplasmy is associated with ß-cell function but not with insulin sensitivity, glucose tolerance, or readouts of muscle mitochondrial quantity and function in m.3243A>G carriers. Exploratory associations between skeletal muscle mtDNA heteroplasmy and outcomes related to glucose homeostasis (A) or muscle mitochondrial quantity and function (B). For each outcome, linear regression models adjusted for age, sex, body mass index (BMI), and moderate-to-vigorous physical activity level (MVPA) were fitted. Reported β values are unstandardized regression coefficients (change in outcome per 1% higher heteroplasmy), with corresponding P values. Dashed lines and shaded bands represent the covariate-adjusted mean prediction and 95% CI at reference covariates. Individual datapoints reflect the observed heteroplasmy level (x-axis) and the model-adjusted outcome (y-axis).

**Figure S5.**
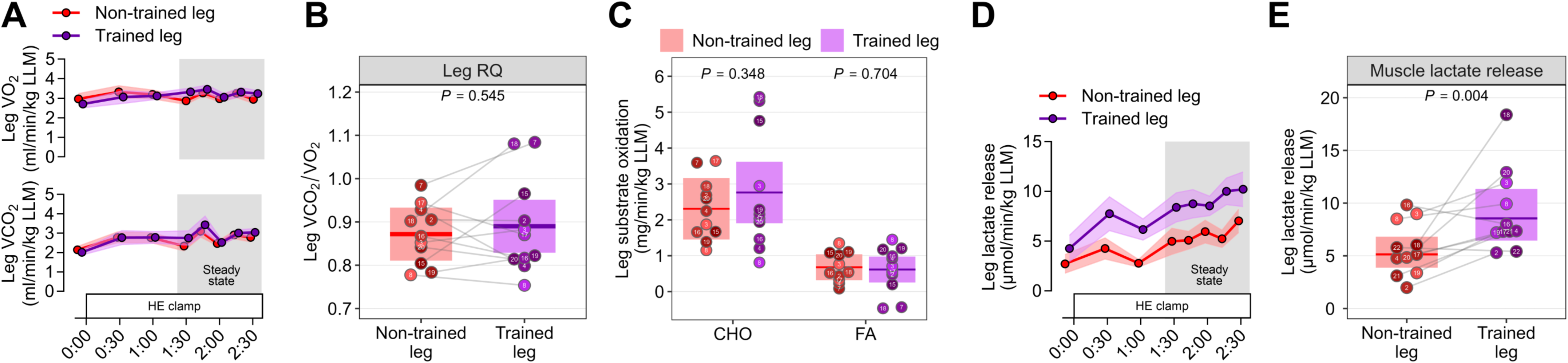
Exercise training does not alter muscle substrate oxidation but increases lactate release in m.3243A>G carriers. (A) Time course of leg O_2_ consumption (V̇O_2_) and CO_2_ release (V̇CO_2_) (data are means ± SEM). (B) Respiratory quotient (RQ) during the steady-state period of the hyperinsulinemic-euglycemic (HE) clamp. (C) Carbohydrate (CHO) and fatty acid (FA) oxidation across the leg, as estimated from the leg V̇O_2_ and V̇CO_2_, during the steady-state period of the HE clamp. (D and E) Leg lactate release as calculated from the arteriovenous difference in plasma lactate concentration and the femoral arterial blood flow. [(D)] Time course of leg lactate release (data are means ± SEM). [(E)]) Leg lactate release during the steady-state period of the HE clamp. Linear mixed models were used to estimate between-leg differences [(B), (C), and (E)]. Data are presented as observed individual values (with lines connecting the non-trained and trained leg from the same participant) and estimated means ± 95% confidence limits. Individual datapoints are color-shaded to indicate muscle mtDNA heteroplasmy (light = low, dark = high). n = 22.

**Figure S6.**
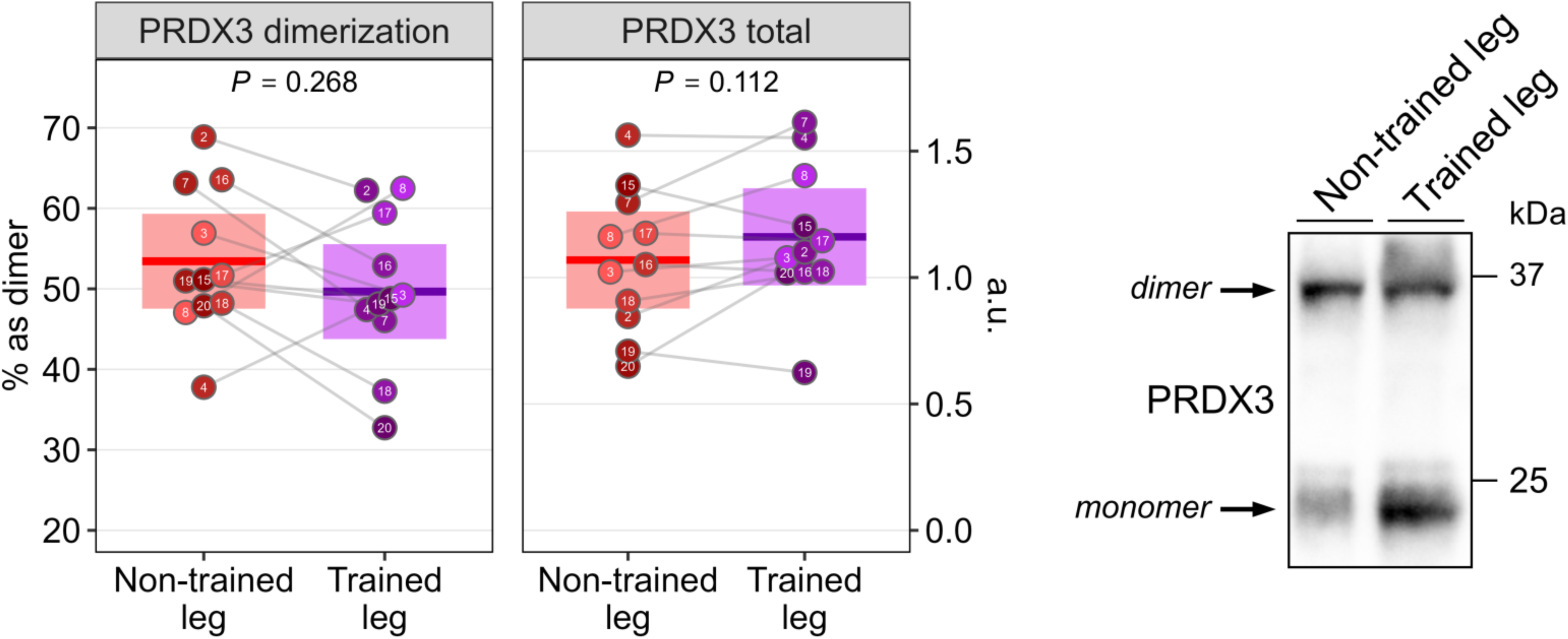
Exercise training does not alter the muscle mitochondrial redox state in m.3243A>G carriers. Mitochondrial redox state as determined by protein abundance of peroxiredoxin 3 (PRDX3) dimers relative to monomers. Linear mixed models were used to estimate between-leg differences. Data are presented as observed individual values (with lines connecting the non-trained and trained leg from the same participant) and estimated means ± 95% confidence limits. Individual datapoints are color-shaded to indicate muscle mtDNA heteroplasmy (light = low, dark = high). n = 22.

**Figure S7.**
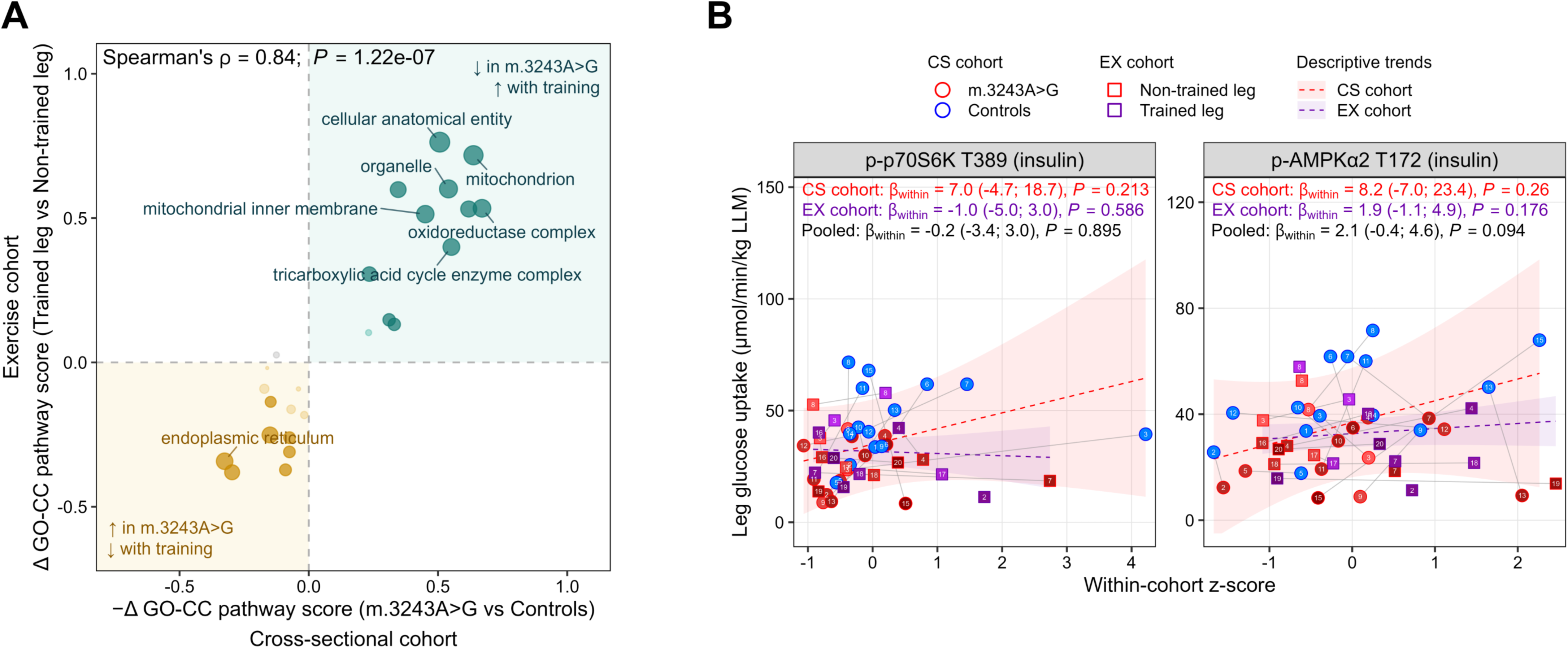
Related to Figure 8. (A) Cross-study integration of m.3243A>G variant defects (cross-sectional cohort, x-axis) and exercise effects (exercise cohort, y-axis) on GO Cellular Component pathway signatures. The scatter plot displays the negated cross-sectional effect (−Δ pathway or protein score; m.3243A>G vs Controls) against the exercise effect (Δ pathway or protein score; trained leg vs non-trained leg), such that features in the upper-right quadrant were suppressed in carriers and rescued by exercise, and features in the lower-left quadrant were elevated in carriers and reduced by exercise. Point size reflects the rescue significance (−log_10_ *P* rescue). Spearman’s ρ and *P* values are shown. Teal points: pathways downregulated in carriers and upregulated by exercise; dark teal: FDR<0.05. Gold points: pathways upregulated in carriers and downregulated by exercise; dark gold: FDR<0.05. (B) Associations between insulin-stimulated mTORC1 and AMPK activation and leg glucose uptake in the cross-sectional (CS) and exercise (EX) cohorts. Analyses were conducted within-clusters, defined as matched pairs (m.3243A>G vs Controls) in the CS cohort or as participants (trained vs non-trained leg) in the EX cohort; the reported β_within_ coefficients reflect covariation within these clusters. Phosphorylated protein abundance was standardized within cohort (z-scored) and a fixed-effects model including cluster as a covariate was fitted separately per study. Cohort-specific β_within_ values represent the change in leg glucose uptake per 1-SD within-cohort increase in the predictor within a cluster, with 95% CI. Pooled β_within_ values were obtained by inverse-variance fixed-effect meta-analysis of the two study-specific estimates. Dashed lines and shaded bands represent cohort-specific marginal trend lines and 95% CI derived from the corresponding fixed-effects models. Thin grey lines connect observations within clusters.

**Figure S8.**
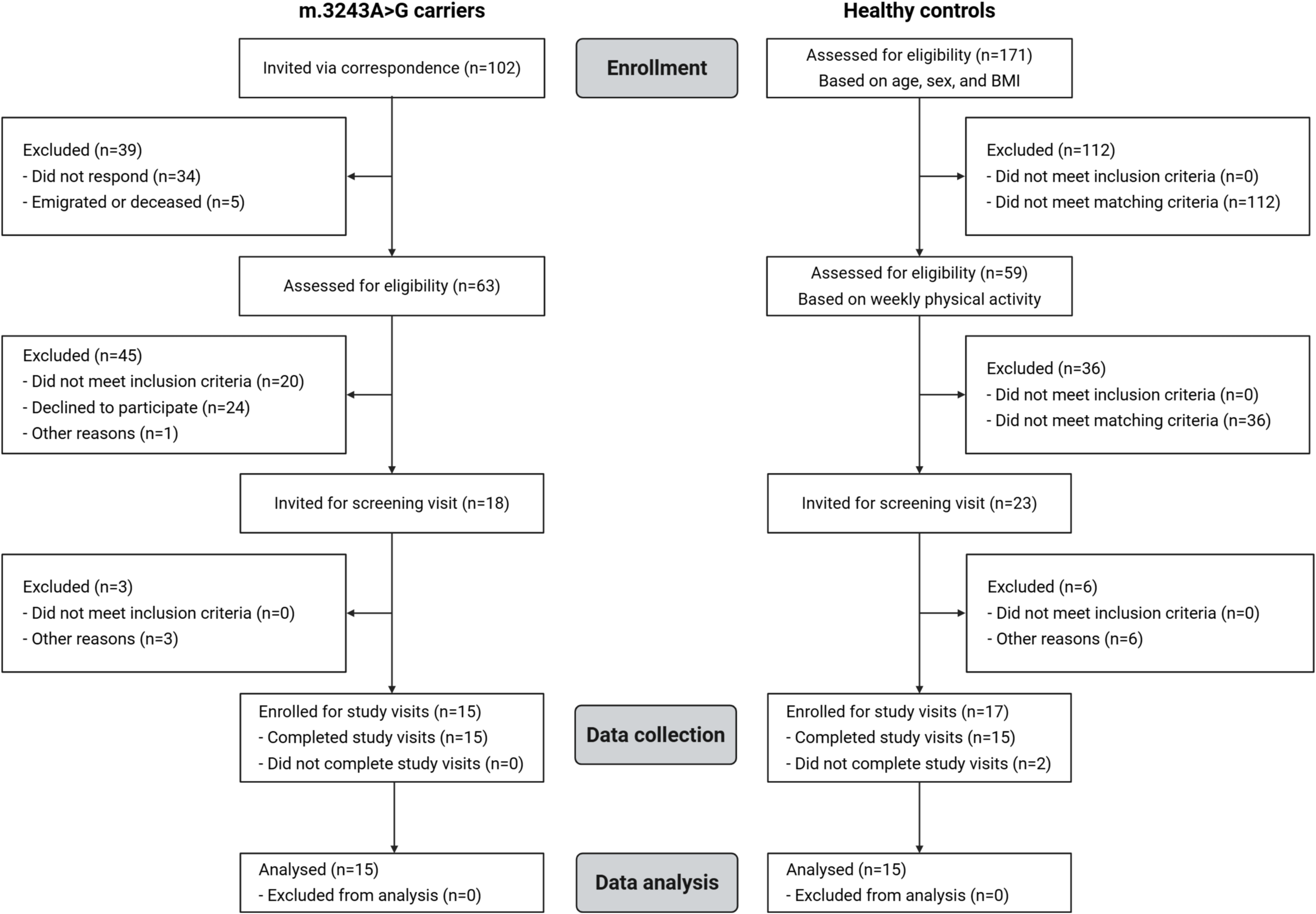
Cross-sectional study flow diagram. Created with BioRender.com

---

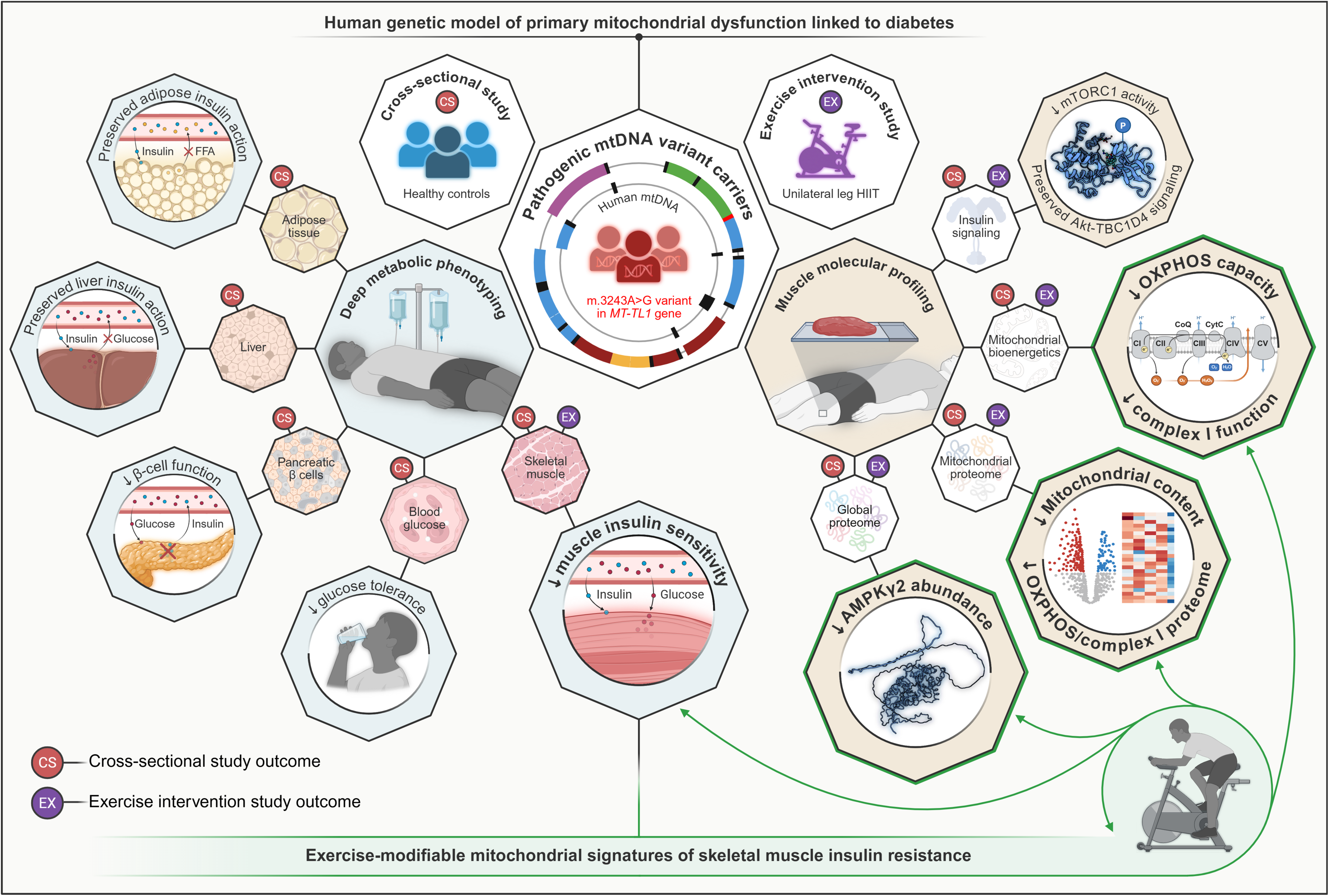

